# Exploring associations between psychotic experiences and structural brain age: a population-based study in late adolescence

**DOI:** 10.1101/2024.10.07.24314890

**Authors:** Constantinos Constantinides, Doretta Caramaschi, Stanley Zammit, Tom P Freeman, Esther Walton

**Affiliations:** Department of Psychology, University of Bath, UK; Department of Psychology, Faculty of Health and Life Sciences, University of Exeter, UK; MRC Centre for Neuropsychiatric Genetics and Genomics, Division of Psychological Medicine and Clinical Neurosciences, Cardiff University School of Medicine, Cardiff, UK; Centre for Academic Mental Health, Population Health Sciences, Bristol Medical School, University of Bristol, Bristol, UK; Addiction and Mental Health Group (AIM), Department of Psychology, University of Bath, UK

**Keywords:** structural brain development, brain age, psychotic-like symptoms, ALSPAC, MRI

## Abstract

Neuroimaging studies show advanced structural “brain age” in schizophrenia and related psychotic disorders, potentially reflecting aberrant brain ageing or maturation. The extent to which altered brain age is associated with subthreshold psychotic experiences (PE) in youth remains unclear.

We investigated the association between PE and brain-predicted age difference (brain-PAD) in late adolescence using a population-based sample of 117 participants with PE and 115 without PE (aged 19-21 years) from the Avon Longitudinal Study of Parents and Children. Brain-PAD was estimated using a publicly available machine learning model previously trained on a combination of region-wise T1-weighted grey-matter measures.

We found little evidence for an association between PEs and brain-PAD after adjusting for age and sex (Cohen’s d = -0.21 [95% CI -0.47, 0.05], p = 0.11). While there was some evidence for lower brain-PAD in those with PEs relative to those without PEs after additionally adjusting for parental social class (Cohen’s *d* = -0.31 [95% CI -0.58, -0.03], p = 0.031) or birth weight (Cohen’s *d* = -0.29 [95% CI -0.55, -0.03], p = 0.038), adjusting for maternal education or childhood IQ did not alter the primary results.

These findings do not support the notion of advanced brain age in older adolescents with PEs. However, they weakly suggest there might be a younger-looking brain in those individuals, indicative of subtle delays in structural brain maturation. Future studies with larger samples covering a wider age range and multimodal measures could further investigate brain age as a marker of psychotic experiences in youth.

## 1. Introduction

While delusions and hallucinations are a key feature of psychotic disorders, these phenomena are also prevalent in the general population, often manifesting at levels that may not reach clinical thresholds and are commonly referred to as psychotic experiences (PEs).^1^ PEs are most common in childhood and adolescence, a critical period of neurodevelopment, and are frequently transient in nature.^2^ Despite their tendency to remit over time, PEs can become persistent and are associated with an increased risk of transition to clinical psychosis.^2–4^ Their manifestation may reflect an atypical developmental trajectory representing vulnerability to psychosis, which in turn may be influenced by environmental and genetic risk factors,^5^ and can be considered as part of a continuum with (ultra) high-risk states and psychotic disorders.^1^ Neuroimaging studies suggest subtle neuroanatomical differences in adolescents with PEs (relative to controls), including global and regional differences in grey matter volume,^6–8^ white matter integrity,^9^ and cortical gyrification.^10,11^ The extent to which apparent structural brain abnormalities associated with PEs reflect atypical brain development or maturation remains elusive.

Using structural magnetic resonance imaging, it is possible to estimate the underlying “biological” age of the brain via supervised machine learning.^12^ The so-called “brain age” paradigm can be seen as a multivariate approach by which advanced algorithms learn age-related, brain-wide structural patterns within normative (training) dataset, and can subsequently be applied to unseen datasets to estimate an individual’s age.^12,13^ Brain-predicted age (or brain age) can differ from actual chronological age, and the discrepancy between the two is captured by the brain-predicted age difference (brain-PAD; also known as brain age gap). While the interpretation of brain-PAD is complex,^14^ a brain-PAD greater than zero indicates a brain that appears ‘older’ than the person’s chronological age and thus may be interpreted as ‘advanced’ brain ageing or maturation, whereas a brain-PAD lower than zero reflects a brain ‘younger’ than expected at a given chronological age (i.e., ‘delayed’ brain ageing or maturation).^13,15^ Large-scale studies covering the adult lifespan have shown a greater brain-PAD in adults with schizophrenia and other psychotic disorders relative to healthy controls (mean brain-PAD ∼ +3.5 years; Cohen’s *d* ∼ +0.5).^16,17^ A greater brain-PAD score has also been observed (albeit to a lesser extent) in adolescents and young adults with schizophrenia or first-episode psychosis^18,19^ and clinical high-risk groups^20,21^. Furthermore, longitudinal data indicate that this gap widens predominantly during the first few years after illness onset before stabilising.^22,23^ While these findings suggest that differences between brain age and chronological age are present early in the course of psychosis, possibly reflecting deviations from a typical neuromaturation trajectory, the developmental period at which these discrepancies emerge and whether they are evident in young people with (subclinical) PEs remain unclear.

In the current study, we aimed to examine whether brain-PAD is associated with PE by utilising data from a population-based case-control sample of older adolescents (19-21 years). We hypothesised a greater brain-PAD score in those with PE (versus those without), and that brain-PAD would be associated with severity of PE, but to a lesser extent than the case-control differences previously observed with clinically ascertained samples. One key advantage of this approach is that it allows examination of PEs in the absence of secondary illness effects such as functional decline or medication. In addition, we explored the association between recurrency of PEs and brain-PAD using longitudinal PE data covering a period from early to late adolescence.

## 2. Methods

### 2.1. Study population and PE classification

We used data from the Avon longitudinal Study of Parents and Children-Psychotic Experiences (ALSPAC-PE) study, which was previously established to investigate the relationship between PEs and brain structure or function.^6,24^ This study is nested within ALSPAC, a population-based cohort established to identify factors influencing child health and developmental outcomes.^25–27^ A more detailed description of participant recruitment can be found in Supplementary section 1 and elsewhere.^6,24^ For the current analyses, individuals were classified as ‘definite, clinical’ PE if they had at least one definite psychotic experience not attributable to sleep or fever that either caused severe distress or had a very negative impact on their social/occupational life or led them to seek help from a professional source. Twenty participants were excluded due to failed image processing or quality control (see Supplementary section 2 for more details) leaving a total of 117 individuals with PE and 115 without PE for the current analyses (N=232).

### 2.2. Structural image acquisition and processing

Structural MRI scans were acquired for each participant using a 3T GT HDx system at Cardiff University Brain Research Imaging Centre (CUBRIC), Cardiff, UK. In the current study, we relied on the image-derived phenotypes extracted centrally by the researchers involved in the ALSPAC neuroimaging resource initiative, which are available via the variable search tool (http://variables.alspac.bris.ac.uk/; and ^24^). Further details on image processing and quality control can be found in Supplementary section 2, Table S1 and the relevant data note by Sharp et al., 2020.^24^

### 2.3. Brain age estimation

To predict brain age in the current sample we used two publicly available models for brain age estimation. First, we used the ENIGMA brain-age model (https://www.photon-ai.com/enigma_brainage)^28^ as it has previously demonstrated relatively good model generalizability to independent cohorts (or scanners) across adulthood as well as the ability to differentiate healthy controls and psychiatric patients (e.g., schizophrenia, major depressive disorder) at the group level.^17,28–30^ This model was trained separately in 952 male and 1,236 female healthy controls aged 18–75 years from the ENIGMA-MDD consortium, using ridge regression.

Recent methodological reports indicate that substantial differences in age distribution between the training and testing samples could potentially negatively affect model generalizability.^31,32^ Therefore, we also applied a second, more recent brain age model developed by the CentileBrain team with a training set that more closely matches the mean age of the current sample (https://centilebrain.org/#/brainAge2).^33^ Briefly, the CentileBrain model was trained separately in healthy females (N=9,185) and males (N=8,328) aged 5-40 years from multiple cohorts from the ENIGMA-Lifespan consortium.

The parameters of the pre-trained sex-specific brain age model(s) were applied individually to each participant within the current sample. Importantly, the current sample was not included in the training sets for either of the two models. Global (i.e., whole-brain) brain-PAD was then calculated for each participant by subtracting chronological age from estimated brain age (i.e., brain-predicted age minus chronological age).

To assess model generalisation performance in terms of age-prediction accuracy, we calculated the mean absolute error (MAE) and a ‘weighted’ MAE (wMAE = MAE / max. – min. age in the training set), taking into account that each of the two models was trained on a different age range. We also report Pearson’s correlation coefficient for brain-predicted age and chronological age (*r*) and explained variance in chronological by brain-predicted age (*R*^*2*^). Further details can be found in Supplementary section 3.

Although we had no pre-specified criteria to reject a model from subsequent statistical analyses, the results obtained with the model showing better age-prediction accuracy (i.e. lowest MAE, wMAE; CentileBrain) constitute our primary analyses, whereas the results obtained with the other model (ENIGMA) are reported as a supplementary analysis.

### 2.4. Non-imaging variables

A small number of early-life factors previously shown to be associated with risk for psychosis were considered in the current study, including parental socioeconomic status,^34,35^ birth weight,^36^ and childhood IQ^37^. The relevant ALSPAC variables included two proxies of parental socioeconomic status (highest level of maternal educational qualification at birth and parental social class), birth weight and childhood IQ at age 8 years. These variables were chosen partly based on data availability for the majority of individuals in the current sample, with missing data ranging from 6% to 13%. For further information, see Supplementary section 4.

### 2.5. Statistical analyses

We used multivariable linear regression to assess the relationship between PE and brain-PAD. In the first model, PE status was specified as the binary predictor of interest (i.e., PE vs. no PEs). In the second model, PEs status was defined as a 4-point ordinal scale (‘none’ > ‘ suspected’ > ‘definite’ > ‘definite, clinical’), as this could potentially provide additional information in terms of PE severity.^6^ We added sex and chronological age as covariates. For further details on brain age bias adjustment, see Supplementary section 5.

A sensitivity power analysis indicated that the current sample (N=232) provides 80% power to detect a relatively small-to-medium standardised effect size (Cohen’s *f* ^*2*^ = 0.034; Cohen’s *d* = ± 0.37) at the typical alpha level of 0.05 (see Supplementary section 6 for more details on power analysis and expected effect size). For details on our sensitivity analyses, see Supplementary section 7. All analyses were performed in R (v. 4.3.0).

## 3. Results

### 3.1. Sample characteristics

#### PE at age 18

The current sample consisted of 117 individuals with PE and 115 without PE as rated at age 18 and were approximately 20 years old at the time of brain scanning (see Table 1). Among those with PEs since age 12, there were 41 participants with suspected PE, 46 with definite (non-clinical) PEs, and 30 with definite, clinical PEs. The latter group had approximately twice the sum of psychotic experiences (median = 2.5) than the other two sub-groups (median = 1; p < 0.0001). Most participants were females (73% and 63% in those with- and without-PE, respectively) and mean age differed slightly across sub-groups. As expected based on previous work with the original imaging sample,^6^ those with PEs had lower maternal education, parental social class, birth weight and childhood IQ relative to those without PEs (Table 1).

**Table 1.**
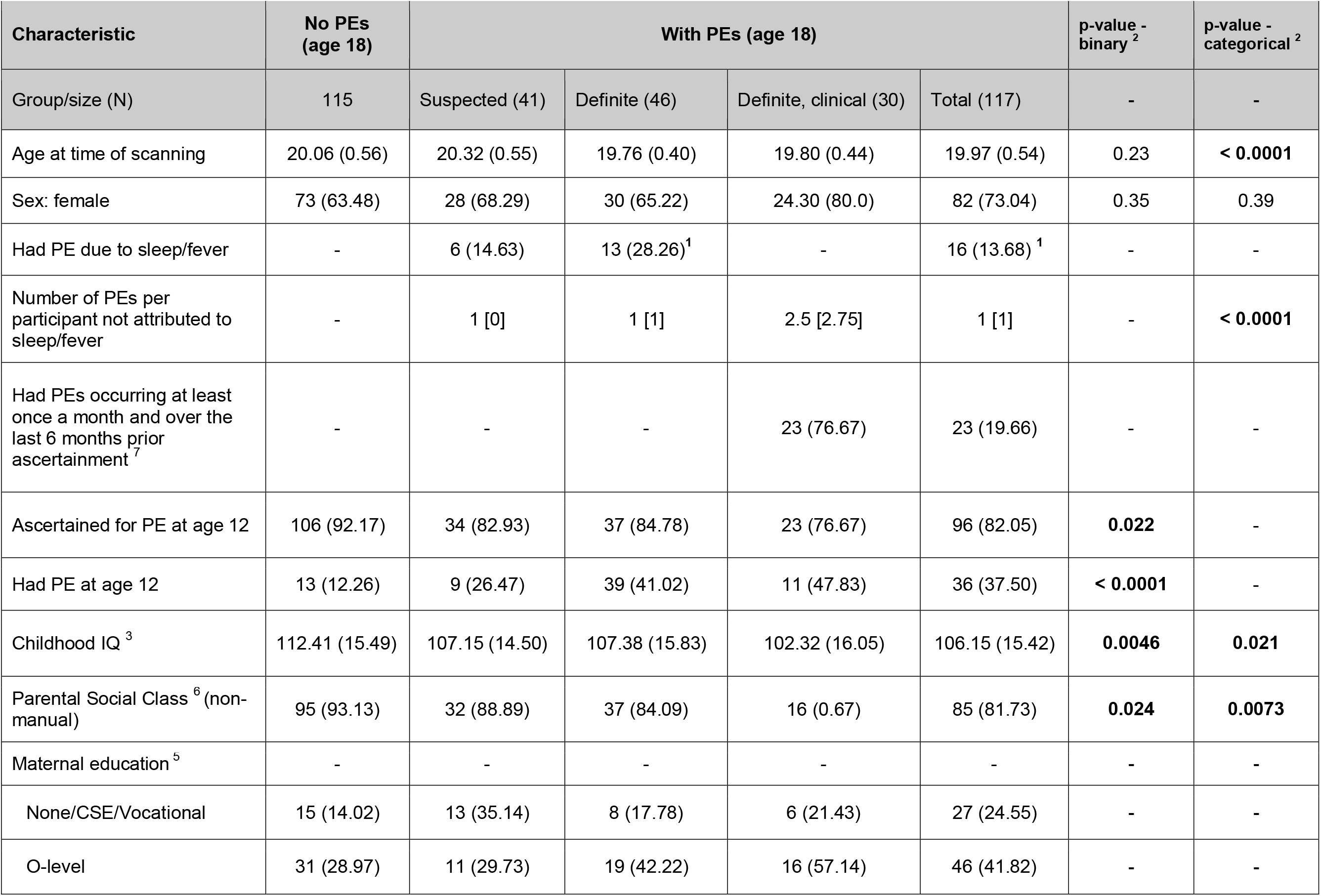
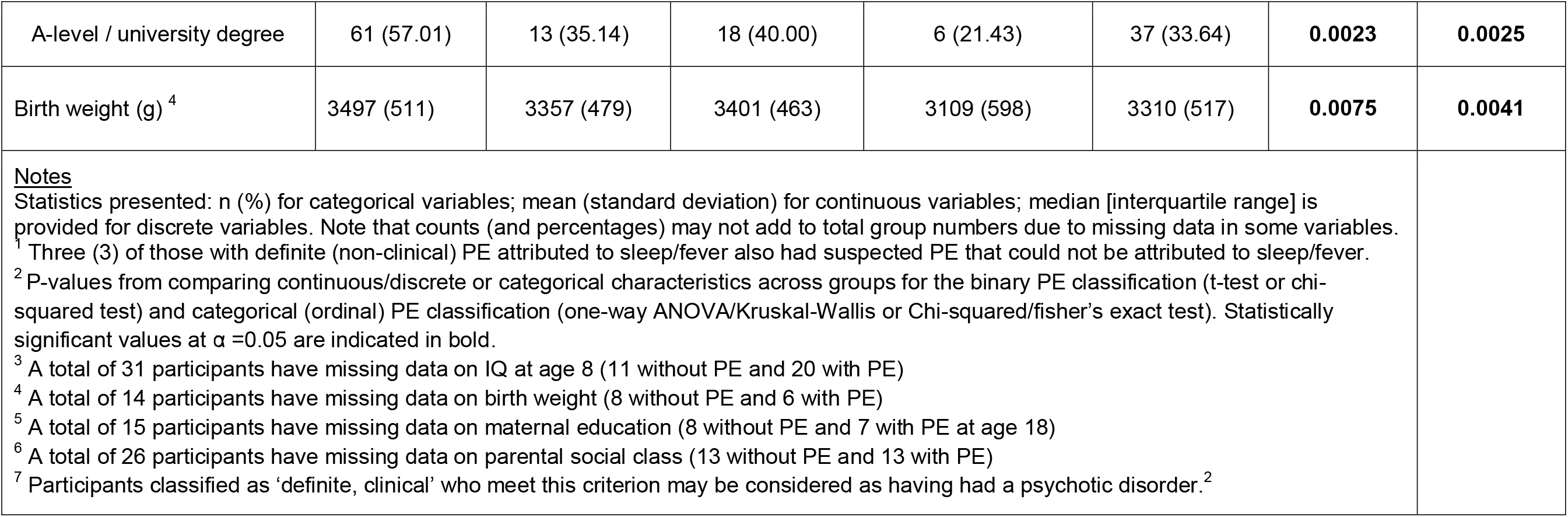
Sample characteristics.

#### Recurrent and transient PE between age 12 and 18

Among those with- and without PE as rated at age 18, 106 (92%) and 96 (82%) had participated in a previous PE assessment at age ∼ 12, respectively (p = 0.002). Within this subset of participants, 36 had recurring PEs, 73 had transient PEs (i.e. 13 at age ∼12 only and 60 at age ∼18 only), and 93 were rated as having no PEs at both time points of ascertainment. Those who were rated as having had PE at age 18 were more likely to also have had PE at age ∼12 (38%) as compared to those without PE (12%; p < 0.0001; Table 1). This observation is consistent with a prior finding in the larger non-imaging study from which the current sample was sourced.^2^

### 3.2. Brain age prediction performance

Despite the very narrow age range of the current sample (19-21 years), there was considerable variation in brain-predicted age as obtained by the CentileBrain model (mean [SD] = 19.41 [2.94] years) and ENIGMA model (24.63 [6.35] years; see Figure 1A). Regardless of PE status, the CentileBrain brain-age model more accurately predicted chronological age with a MAE of 2.36 years (SE = 0.19, wMAE = 0.067, r = 0.13) in males and 2.47 years (SE = 0.15; wMAE = 0.071, r = 0.006) in females (Fig. 1B). Although age-prediction accuracy is not directly comparable between different studies,^31^ these errors are in good alignment with those reported in previous studies of brain age in adolescence using external validation sets (overall age range: 5-22 years; MAE: 0.7-2 years).^19,38–40^ On the other hand, the ENIGMA model moderately predicted chronological age with a MAE of 5.43 years (SE = 0.48, wMAE = 0.095, *r* = 0.11) for males and 6.83 years (SE = 0.36, wMAE = 0.12, *r* = 0.06) for females (Fig. 1C). Post-hoc t-tests indicated a significantly lower age-prediction accuracy in females compared to males for the ENIGMA model (mean difference in MAE = +1.4 years; p=0.022), but not for the CentileBrain model (mean difference = +0.10 years; p=0.67). Performance metrics followed a largely similar trend when calculated separately in those with or without PEs (see Supplementary Table S2).

**Figure 1.**
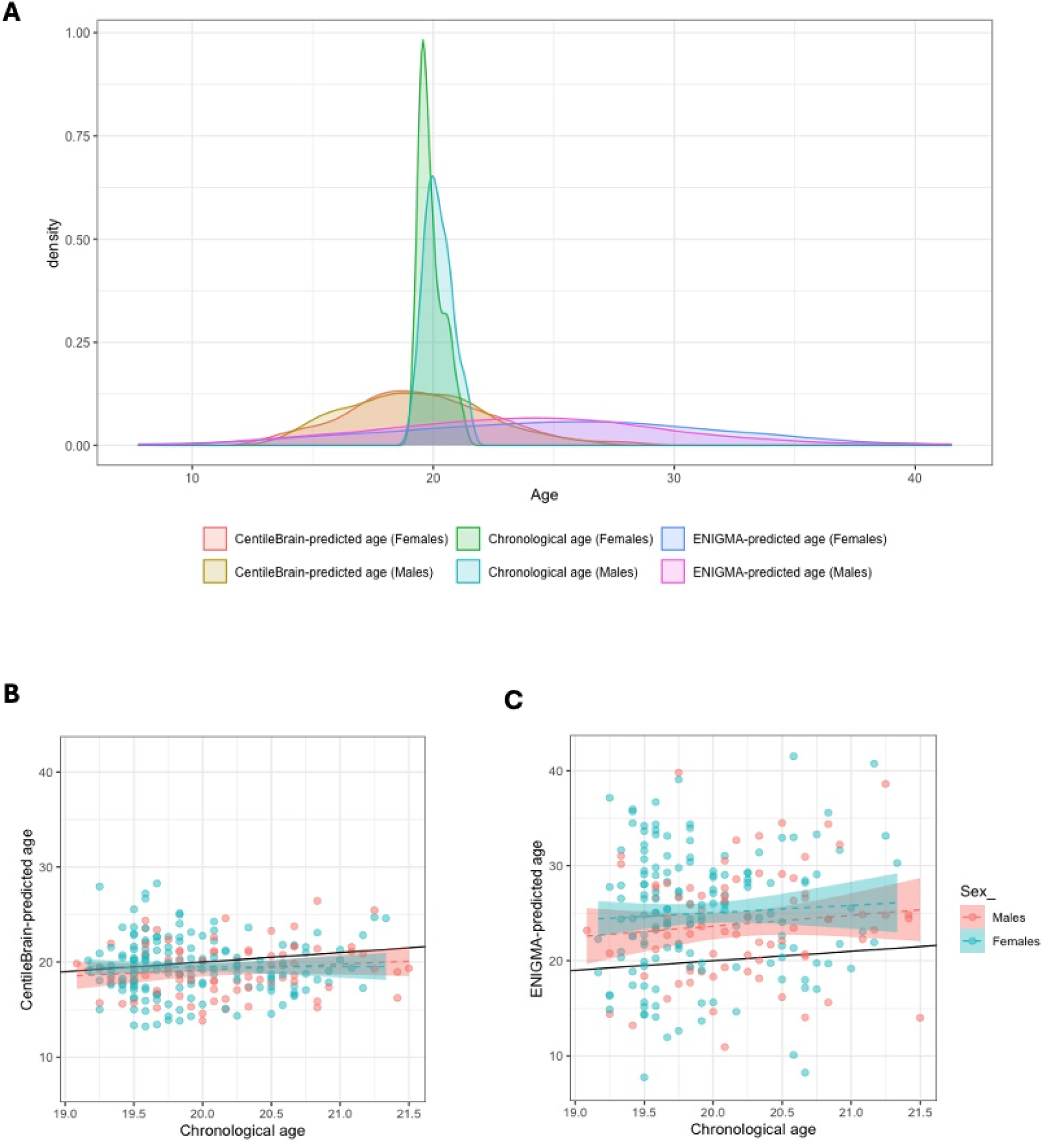
Chronological age and brain-predicted age in the current sample. **(A)** Density plots of brain-predicted age (years) for the sex-specific CentileBrain and ENIGMA models and with reference to chronological age distribution. **(B)** Chronological age versus brain-predicted age for the CentileBrain model. The black line represents an (hypothetical) exact linear relationship between chronological age and brain-predicted age (years). Dashed lines represent the actual fit (with standard error shading) between chronological and brain-predicted age for each sex. A tendency toward underestimation of brain-predicted age for relatively older participants (> ∼ 20 years) was observed in both sex groups. **(C)** Similar to (B), chronological age versus brain-predicted age for the ENIGMA model. A considerable overestimation of brain-predicted age across chronological age was observed in both sex groups.

In terms of age-related bias, the CentileBrain model systematically underestimated brain age particularly among relatively older participants of the current sample regardless of sex (Fig. 1B) or PE status (Fig. S1), with a negative albeit weak linear dependence of brain-PAD on chronological age (*r* = -0.15; p=0.03; mean brain-PAD = -0.60 years; Fig. S2). As noted in section 2.4, this was dealt by adjusting for chronological age in subsequent analyses. Brain age was considerably overestimated by the ENIGMA model across participants of the current sample (mean brain-PAD = +4.62 years), regardless of sex (Fig. 1C) or PE status (Fig. S1), with no observed linear dependence of brain-PAD on chronological age (*r* = -0.03; p=0.62; Figure S2). Nonetheless, age was added as a covariate in subsequent analyses to account for shared variance between predictors. CentileBrain-derived brain-PAD and ENIGMA-derived brain-PAD estimates were only moderately correlated with one another (age adjusted *r* = 0.52, p < 0.0001; Figure S3).

The rest of this section summarises the results obtained with CentileBrain-derived brain-PAD as the primary outcome measure (sections 3.3 to 3.6), whereas results obtained with ENIGMA-derived brain-PAD are reported as a supplementary analysis (see supplementary materials).

### 3.3. PEs and brain age

Mean brain-PAD was -0.86 (SD = 3.15) years in those with PEs and -0.34 years (SD = 2.75) years in those without PEs at age 18. There was a weakly negative albeit non-significant difference in mean brain-PAD between the those with and without PEs, adjusting for age and sex (b = -0.62 [SE = 0.39] years; Cohen’s *d* = -0.21 [95% CI -0.47, 0.05], p = 0.11; Figure 2A). With respect to the ordinal PE classification, there was a weakly negative but non-significant association with brain-PAD (b = -0.26 [SE=0.18] years, partial *R*^*2*^ =0.009, p = 0.11; see Fig. 2B). Post-hoc analyses provided no evidence for a linear trend across the PE sub-groups, however the mean difference in brain-PAD between each PE group (suspected/definite/definite, clinical) and controls was in the negative direction (Figure S4).

**Figure 2.**
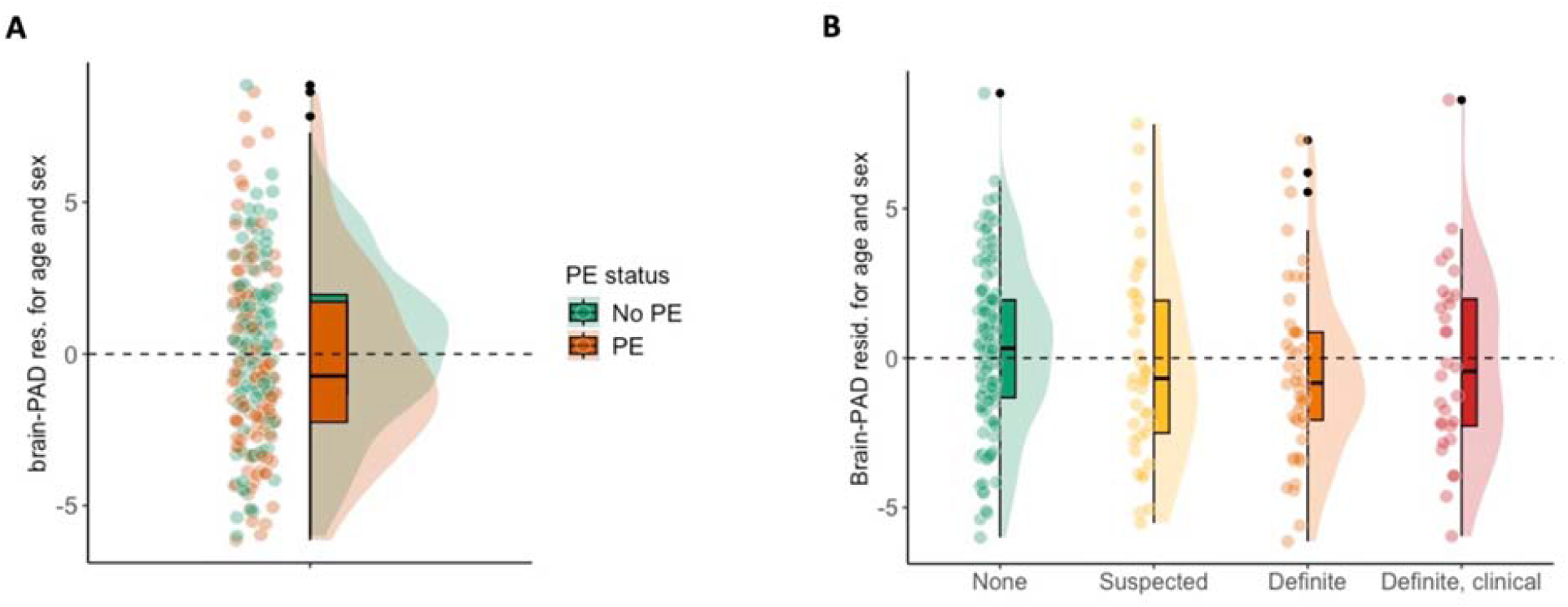
Psychotic experiences and brain-PAD. **(A)** brain-PAD estimates among participants with and without PE (binary classification). **(B)** Brain-PAD estimates among those without PE (none) and each PE sub-category (suspected, definite (non-clinical), definite, clinical). Plotted brain-PAD estimates were residualised for age and sex. Raincloud (half-density) plots depict group median, interquartile range, and potential outliers.

### 3.4. PE recurrence and brain age

Analysis of compete-case longitudinal PE data across age 12 and age 18 years did not support an association between recurring-or transient-PEs and brain-PAD, however effect estimates were fairly consistent with those observed for PEs at age 18 in terms of size and direction (*recurring* PEs: b = -0.61 [SE=0.56] years, Cohen’s *d* = -0.18 [95% CI -0.56, 0.21], p=0.27; *transient* PEs: b = -0.56 [SE=0.44] years, Cohen’s *d* = -0.18 [95% CI -0.49, 0.12], p= 0.20).

### 3.5. Sensitivity analyses

Repeating the above-described analyses after excluding a small number of brain-PAD outliers (n=2/1; Tables S3-S5 and Figure S4) or reclassifying those who solely had PEs attributed to sleep/fever (n=13) to controls (Table S6), led to highly comparable results. The results of the primary analyses were also robust to further adjustment for each of the potentially confounding early-life risk factors considered (Table S7). Adjusting for parental social class led to a significant albeit weakly negative association between PEs and brain-PAD (*binary* PE: b = -0.88 years [SE=0.40], Cohen’s *d* = -0.31 [95% CI -0.58, -0.03], p = 0.031; *ordinal* PE: b = -0.43 years [SE=0.19], partial *R*^*2*^ = 0.025, p = 0.024). Adjusting separately for birth weight also provided some limited evidence for a negative association (*binary* PE: b = -0.85 years [SE=0.40], Cohen’s *d* = -0.29 [95% CI -0.55, -0.03], p = 0.038, *ordinal* PE: b = -0.35 years [SE=0.19], partial R2 = 0.016, p = 0.066).

### 3.6. Correlations between Freesurfer features and brain-predicted age

Most FreeSurfer features were negatively correlated to brain-predicted age at varying degrees across the total sample, with a small number of features such as right hippocampal volume and right parahippocampal thickness showing relatively weak positive correlations (overall Pearson’s *r* range: -0.41 to +0.15; Table S8). On average, cortical thickness features showed relatively stronger negative correlations (mean *r* [SD] = -0.18 [0.13]) than cortical surface area features (−0.12 [0.07]) and subcortical grey matter volumes (−0.11 [0.11]). With respect to individual brain regions, the top correlations in terms of magnitude were located in thickness of left and right precuneal, posterior cingulate, superior frontal, superior parietal, inferior parietal and rostral middle frontal cortices, and right paracentral cortex (*r* range: -0.28 to -0.41). The relative strength of these top correlations were fairly comparable within the control (no-PE) and PE groups (Table S8).

## 4. Discussion

We investigated the association between a putative biomarker of brain maturation (or “ageing”) and psychotic experiences (PEs) in a population-based sample of older adolescents. Contrary to our expectations, we found little evidence to suggest an association between PEs and brain-PAD.

The results of the current study should be interpreted in context with the existing literature. Using a community-based sample of youths aged 8-21 years, Cropley et al (2021)^41^ found a higher, albeit small increase in brain-PAD (∼ +0.15 years; *d* ∼ +0.18) in those endorsing (subclinical) psychotic symptoms (n=328) and their typically developing peers (n=402). Moreover, they found a group-by-age interaction indicating that this overall positive group difference was driven by younger (< ∼ 16 years) rather than older adolescents (17-21 years).^41^ Thus, the discrepancy in effect size direction with regard to the current study (d = -0.21) could be attributed to differences in overall age range between the two studies. Chung et al (2021)^20^ investigated brain age in youths with clinically high risk for psychosis (n=275) and found a higher brain-PAD (+0.64 years) relative to healthy controls (n=109; aged 12-21 years). However, this association was moderated by chronological age, with a positive brain-PAD difference present in younger adolescents who developed psychosis (12-17 years), and not in older adolescents (18-21 years). It should also be emphasised that the current analyses were not adequately powered to detect small effect sizes such the one observed in our primary analyses (*d* = -0.21), thus our results should be interpreted with caution.

Overall, the current study is inconclusive about the association between PEs and brain-PAD, with some evidence for a between-group difference only after adjusting for the potentially confounding effects of birth weight or parental socioeconomic status. Nevertheless, it is interesting that the observed effect size lies in the opposite direction (i.e. negative) than the one we primarily hypothesised based on prior work in young people with a psychotic disorder or at clinically high risk for psychosis.^19–21^ A more negative brain-PAD score among older adolescents with PEs within the current sample may reflect “delayed” structural brain maturation. This interpretation aligns with a finding from a previous analysis of this sample by Drakesmith et al (2016)^6^ reporting increasing severity of PEs to be associated with a reduction in T1 relaxation rate across right prefrontal and left temporoparietal cortices, with the latter having been previously identified to exhibit a relatively prolonged period of cortical development during adolescence.^6,42^ While these cortical areas may partly overlap with the imaging feature-brain age correlations reported here, it is important to emphasise that those correlations may primarily reflect the relative contribution of those features in brain age-prediction rather than the observed between-group differences in brain-PAD. Despite this limitation, individual brain-PAD scores were estimated based on a very large and healthy (training) sample covering critical developmental periods from childhood through early adulthood (CentileBrain model). As such, the current analyses may reinforce these prior developmental inferences regardless of the cross-sectional nature and narrow age range of this sample (19-21 years). Nonetheless, without longitudinal imaging data the interpretation of a negative brain-PAD score remains uncertain. For example, it may reflect earlier disturbances in brain development or integrity,^14^ rather than a deviation from a typical brain maturation trajectory during adolescence. As the observed association between PEs on brain-PAD was sensitive to adjustment for birth weight in the current study, this interpretation warrants some consideration. An early disturbance in brain development is to some extent supported by the work of Fonville et al, 2019^10^ using this sample, showing an association between recurring PEs and reduction in local gyrification index (a measure of cortical folding) in left temporal gyrus, which appeared to have occurred in isolation from white matter changes that typically take place during late adolescence. However, a more recent longitudinal large-scale study by the IMAGEN consortium provided evidence for “dynamic” gyrification changes as reflected by higher cortical gyrification in the right parietal cortex in older adolescents with elevated levels of PEs (and since early adolescence), in addition to “static” changes at fronto-temporal regions in the left hemisphere.^11^ This may suggest that a more negative brain-PAD in those with PEs as observed in the current study could in part reflect altered brain maturation during adolescence that is more proximal to manifestation of psychotic symptoms, regardless of their temporal relationship.

The current study has some further limitations that need to be taken into consideration. First, while psychotic experiences were qualitatively ascertained based on semi-structured interview (PLIKS), thus more closely approximating clinical ascertainment than self-reported measures,^2^ the ordinal PE variable does not constitute a truly dimensional measure of psychotic experiences. This might have limited its ability to explain potentially relevant variance in brain-PAD. Furthermore, the additional analyses for association between recurring or transient PEs and brain-PAD are limited by an even smaller sample (due to missing data at age 12) and incorporating PE severity in those analyses was not suitable due to lower sub-group numbers. Second, while the narrow age range (19-21 years) might have helped minimise the effects of temporal characteristics of sexual development, such as pubertal status in early adolescence,^39^ any inferences that could be drawn from the current study may be limited to late adolescence. Third, both models used to estimate brain age were based solely on T1-weighted MRI with grey-matter features derived from a low-dimensional cortical atlas. As brain maturation (or ageing) is a heterogeneous process, future studies could employ brain-age measures based on higher dimensionality sMRI data and/or other imaging modalities to potentially increase precision in brain age estimation or sensitivity to individual differences.^43–45^ Lastly, and as noted above, the current study was focused on a single “global” measure of brain age that could overlook any localised (or brain region-specific) deviations in brain maturation as linked to PEs. While this is a common limitation of brain age studies, future work could consider brain age estimation both at the global as well as local level.^46,47^

In summary, the current study did not find strong evidence for an association between PE and (global) structural brain age in older adolescents. However, the results weakly suggest there might be a younger-looking brain in those individuals (negative brain-PAD), indicative of subtle delays in structural brain maturation. Future studies with larger samples, longitudinal designs, and more comprehensive measures could further explore the relationship between PEs and this putative marker of structural brain maturation.

## Supporting information

Supplementary section

## Data Availability

The authors do not have permission to share data. Researchers can request the original dataset used directly from ALSPAC (https://www.bristol.ac.uk/alspac/researchers/). The ALSPAC website contains details of all the data that is available through a fully searchable data dictionary and variable search tool (https://www.bristol.ac.uk/alspac/researchers/our-data/).

## 5. Author Contributions

*Conceptualization*: CC, EW, TF. *Methodology*: CC, EW, TF, SZ. *Formal analysis*: CC. *Resources*: EW. *Writing -Original Draft*: CC. *Writing - Review & Editing*: CC, EW, TF, DC, SZ. *Supervision*: EW, TF, DC, SZ.

## 6. Acknowledgements

We are extremely grateful to all the families who took part in this study, the midwives for their help in recruiting them, and the whole ALSPAC team, which includes interviewers, computer and laboratory technicians, clerical workers, research scientists, volunteers, managers, receptionists, and nurses. We also thank Dr. Sophia Frangou for providing additional background information on the training and validation of the CentileBrain model.

## 9. Data and code availability

The authors do not have permission to share data. Researchers can request the original dataset used directly from ALSPAC (https://www.bristol.ac.uk/alspac/researchers/). The R code used for the current study will be made available via GitHub (https://github.com/ConstantinosConst/ASLPAC-PE-BrainAge.git).

