## Supplementary section for "Exploring associations between psychotic experiences and structural brain age: a population-based study in late adolescence"

**Supplementary Materials**

**Contents**

**Supplementary Methods and Results**

1. Study population and PE classification
2. Structural image acquisition and processing
3. Brain age estimation
4. Non-imaging variables
5. Brain age bias adjustment
6. Sensitivity power analysis
7. Sensitivity and additional analyses

8. Supplementary analysis with ENIGMA-derived brain-PAD

**Supplementary Tables**

Supplementary Table S1. Basic characteristics of excluded participants

Supplementary Table S2. Brain age model performance in the current sample

Supplementary Table S3. Primary analyses of the association between binary or ordinal PE classification and brain-PAD, before and after exclusion of brain-PAD outliers.

Supplementary Table S4. Sub-group analyses of difference in mean-brain-PAD between each PE category and those without PE, before- and after- exclusion of brain-PAD outliers

Supplementary Table S5. Exploratory analyses of the association between transient or recurring PE and brain-PAD, before and after exclusion of brain-PAD outliers.

Supplementary Table S6. Sensitivity analyses of the association between brain-PAD and PEs not attributable to sleep/fever, before- and after- exclusion of brain-PAD outliers.

Supplementary Table S7. Supplementary Table S7. Sensitivity analyses of the effect PEs on brain-PAD additionally adjusting for potential confounders (risk-factors)

**Supplementary Figures**

Supplementary Figure S1. Chronological versus brain-predicted age with respect to PE status.

Supplementary Figure S2. Age-related bias in brain age prediction

Supplementary Figure S3. Pearson’s and spearman’s correlation between ENIGMA derived brain-PAD and CentileBrain-derived brain-PAD

Supplementary Figure S4. Difference in brain-PAD between suspected, definite, or definite, clinical PEs and those without PEs (reference).

Supplementary Figure S5. Psychotic experiences and ENIGMA-derived brain-PAD

**Supplementary References**

**Supplementary Methods and Results**

1. **Study population and PE classification**

The broader ALSPAC study originally invited pregnant women residing in Avon (South-West England) with expected delivery dates between 1st April 1991 and 31st December 1992. The initial number of pregnancies enrolled was 14,541, resulting in 13,988 children who were alive at 1 year of age. When the oldest children were approximately 7 years of age, an attempt was made to bolster the initial sample with eligible cases who had failed to join the study originally. Consequently, the total sample size for analyses using any data collected after the age of seven is therefore 15,447 pregnancies and 14,901 children who were alive at 1 year of age. The phases of enrolment and study representativeness are described in more detail in the cohort profile paper and its updates.^1–3^

Ethical approval for the study was obtained from the ALSPAC Law and Ethics Committee and the Local Research Ethics Committees (listed at http://www.bristol.ac.uk/alspac/researchers/research-ethics/). Informed consent for the use of data collected via questionnaires and clinics was obtained from participants following the recommendation of the ALSPAC Ethics and Law Committee at the time. The ALSPAC website contains details of all the data that is available through a fully searchable data dictionary and variable search tool (<https://www.bristol.ac.uk/alspac/researchers/our-data/>).

At approximately the age of 18 years, a subsample of 4,320 ALSPAC offspring were screened for psychotic experiences,^4^ using the semi-structured Psychosis-Like Symptoms Interview (PLIKS) as described previously.^5,6^ PLIKS builds upon clinical criteria developed as part of the Schedule for Clinical Assessment in Neuropsychiatry (SCAN). Approximately 10% of the subsample (n=433) were found to have had one or more ‘suspected’ or ‘definite’ psychotic experiences (i.e., hallucinations, delusions, or experiences of thought interference) since age 12.^4^ Of these, 126 were recruited to participate in a neuroimaging sub-study. From the remaining participants who were identified as not having had PEs (n=3,887), an equal number (n=126) were randomly recruited as controls. Participants were between 19 and 21 years old at the time of brain scanning.

A subset of the above-described sample had also participated at a previous PE assessment around the age of 12 (N=202).^5^ We have further classified these participants with respect to recurrence of PEs across the two time points of ascertainment; those who rated as having at least one PE at either point of assessment were classified as ‘transient’ PE, whereas those rated with PE at both assessments were classified as ‘recurring’ PE.

2. **Structural image acquisition and processing**

T _1_-weighted structural images were acquired using a FSPGR sequence (TR = 7.8 ms, TE = 3.0 ms, TI = 450 ms, flip angle = 20°, voxel size = 1 mm^3^ isomorphic resolution)^7^ and processed using FreeSurfer (version 6.0.0) to extract cortical and subcortical measures from multiple regions of interest (ROIs) based on the Desikan-Killiany (DK) atlas and Aseg atlases.^8^ Reconstructed images and their cortical and subcortical parcellations/segmentations underwent quality control following standardised protocols developed by the ENIGMA consortium (<http://enigma.ini.usc.edu/protocols/imaging-protocols/>). Each T1-weighted MRI scan was segmented and parcellated bilaterally into volumes for 7 subcortical grey-matter regions (2 x 7; left and right nucleus accumbens, amygdala, caudate, hippocampus, pallidum, putamen, and thalamus) and 2 lateral ventricles (left and right), 34 regional cortical thickness (left and right; 2 x 34) and cortical surface area (2 x 34) measures, and total intracranial volume (ICV; N_measures_ = 153). Out of 252 participants in the ALSPAC PE sub-study, six had scans that failed image reconstruction and thus were excluded from the current analyses. A further 14 participants were excluded due to failed quality control for cortical parcellation (n=10) or subcortical segmentation (n=4; see Supplementary Table S1 for basic characteristics of excluded participants).

3. **Brain age estimation**

The ENIGMA brain age model was trained separately in 952 male and 1,236 female healthy controls aged 18–75 years from the ENIGMA-MDD consortium, using ridge regression. FreeSurfer measures from the left and right hemispheres were combined by calculating the mean ((left + right)/2)) of volumes for subcortical regions (n=7), lateral ventricles (n=1), and thickness (n=34) and surface area (n=34) for cortical regions, and ICV, resulting in a total of 77 input features for brain age prediction. Within the training set, under cross-validation, the sex-specific models achieved a mean absolute error (MAE) of 6.32 years (*r* = 0.85) in males and 6.59 years (r=0.85) in females.^9^ The ENIGMA model was trained in relatively older samples with mean age [SD] of 43.3 [15.2] and 40.0 [15.7] in males and females, respectively, but our testing sample includes youths aged 19-21 years.

The CentileBrain model was trained separately in healthy females (N=9,185) and males (N=8,328) aged 5-40 years (mean age [SD] of 17.79 [7.63] and 18.24 [7.44] in males and females, respectively) from multiple cohorts from the ENIGMA-Lifespan consortium. The model utilises support vector regression with radial basis function kernel, which was previously shown to demonstrate optimal age-prediction accuracy in using DK atlas features among children and adolescents whilst exhibiting robustness to outliers as well as having the ability to model nonlinear and interactive relationships between brain imaging features and age.^10^ In a fairly similar manner to the ENIGMA model (albeit excluding ICV and lateral ventricles), FreeSurfer features included 34 cortical thickness and 34 cortical surface area regional measures, and 7 subcortical grey matter volumes, per each hemisphere (i.e., 75 x 2 = 150 input features). Under cross-validation within the training set, the sex-specific models achieved a MAE = 2.45 (r = 0.89) and MAE 2.38 (r=0.89) in males and females, respectively.^11^

Global (i.e., whole-brain) brain-PAD was calculated for each participant in the current sample by subtracting chronological age from estimated brain age (i.e., brain-predicted age minus chronological age). Of note, brain-PAD scores do not reflect a unitary biological process regardless of the brain-age model used to derive them. Individual brain-PAD scores may reflect very different mechanisms depending on the individual’s developmental period and/or health condition.^12,13^

To assess model generalisation performance in terms of age-prediction accuracy, we calculated MAE between brain-predicted age and chronological age with respect to sex and/or PE status in the current sample (lower values indicate better fit). To aid comparison between the two brain-age models we have also calculated a ‘weighted’ MAE (wMAE = MAE / max. – min. age in the training set), taking into account that each of the two models was trained on a different age range. This metric is based on prior methodological research demonstrating that MAE inevitably decreases as the age range of the training set becomes narrower,^14^ however wMAE does not take into account the underlying differences in age distribution of the training and testing sets. We also report Pearson’s correlation coefficient for brain-predicted age and chronological age (*r*) and explained variance in chronological by brain-predicted age (*R^2^*); while a higher *r or R^2^* typically indicates a better model fit**,** these metrics should be interpreted with caution given the very narrow age range of the current sample (19-21 years)**,** which inevitably leads to less covariance between brain-predicted age and actual age regardless of prediction accuracy.^14^

4. **Non-imaging variables**

ALSPAC variables included two proxies of parental socioeconomic status - highest level of maternal educational qualification at birth (here categorised as low = none, CSE or vocational; medium = O-level or equivalent; high = A-level or equivalent, or university degree) and parental social class (highest among both parents; social classes were dichotomized as non-manual [I, II and III-non-manual combined], and manual [III-manual, IV, and V combined]).^15,16^ Birth weight as identified through a variety of sources including obstetric data and birth notifications. Childhood IQ at age 8 years was assessed using the short form of Wechsler Intelligence Scale for Children (WISC-III).^17^

**5. Brain age bias adjustment**

There is a well-described age-related bias inherent to the ‘brain age’ prediction framework, where brain age is overestimated in younger individuals and underestimated in older individuals (relative to the age distribution of the training set), and most accurately estimated for individuals with an age closer to the average age (of the training set) that is thought to be due to regression toward the mean.^18–20^ Several bias-adjustment procedures have been developed to account for this chronological age dependency (for an overview, see de Lange & Cole, 2020).^21^ Unless otherwise specified, here we added chronological age as a covariate in our statistical analyses to account for linear relationships between brain-PAD and chronological age.^18^ In addition, individual brain-PAD estimates were residualised for age, where appropriate, for data visualisation only. Importantly, we did not adjust performance metrics (i.e., MAE, *r*) for this age-bias as we chose to focus on raw (out–of-model) performance and because such corrections were previously characterised as potentially misleading.^14,22^

**6. Sensitivity power analysis**

We added chronological age as a covariate in our statistical analyses to account for linear relationships between brain-PAD and chronological age.^18^ In addition to chronological age, sex was added as a covariate in both models to account for independent effects of sex on brain-PAD as shown in previous work with adolescent/young adult samples.^23,24^ We used a two-tailed null hypothesis test (H_0_: β=0) and 95% confidence intervals to make inferences about the association between PE and brain-PAD. Standardised effect size estimates for group differences were calculated using Cohen's *d*, derived from the t-statistic of the binary PE variable from the regression models,^25^ and partial R^2^ for the ordinal PE variable.

*Minimum detectable effect size*

Based on our total sample (N=232), we had 80% power to detect an effect of Cohen’s *f ^2^* = 0.034 (or higher) in terms of the proportion of variance in brain-PAD explained by PE status at the typical alpha level α = 0.05, using linear multivariable regression with a total of 3 predictors (PE status, age, sex). This minimum detectable standardised effect size was estimated with G*Power using the *“linear multiple regression: Fixed Model, R^2^ increase (F-test)”* function and thereafter validated using the *pwr.f2.test* function of the ‘pwr’ package in R (<https://github.com/heliosdrm/pwr>). The minimum detectable Cohen’s *d* for our binary PE model (PEs vs. No PEs) can be derived from the following equation: *f ^2^* = *d* ^2^ / 2k (where k is the number of groups of equal size).^26^ The minimum detectable Cohen’s *d* is therefore ± 0.37, which can be considered as ‘small-to-medium’ effect size.^26^

*Expected effect size*

A previous study by Cropley et al (2021)^27^ reported a relatively weak correlation between brain-PAD and a (sub)clinical psychosis symptoms rating scale (t=3.16; p=0.0016; r = 0.11**;** adjusted for chronological age) in a large, young community-based sample of typically and non-typically developing youths aged 8-21 years (N=1313). When substituted the continuous psychotic psychopathology dimension with a categorical operationalization, there was a positive mean brain-PAD difference between those with lifetime endorsement of subclinical psychotic symptoms (n=328) relative to typically developing youths (n= 402; t= 2.35; p=0.02; adjusted for chronological age).^27^ A standardised effect size (Cohen’s *d*) for this group difference can derived from a general linear model using the following formula:

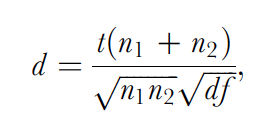

where *t* represents the test statistic (*t* = 2.35) and df represents the regression’s degrees of freedom for the test (= 402+329 - 2 - 1). n1 (=329) and n2 (=402) represent the sample size of each of the two groups.^25^ The derived Cohen’s *d* is + 0.18, which is typically considered as a “small” effect size.^26^ Given the above sensitivity power analysis, this suggests that the current study may not be adequately powered to detect an association between PEs and brain-PAD at the typical alpha level α = 0.05. Nonetheless, it should be emphasised that the brain-age model(s) and tool for ascertaining PE as used in the current study are not necessarily equivalent to those used by Cropley et al (2021),^27^ and the two studies differ in terms of age range (i.e. 19-21 years vs. 8-21 years) and control group definition. Thus, this “expected” effect size (d = +0.18) should be interpreted with caution.

**7. Sensitivity and additional analyses**

*Sensitivity analyses*

As a subset of participants had ‘suspected’ or ‘definite’ PEs solely attributed to sleep or fever (n=16), sensitivity analyses were performed in which those participants were reclassified as having no PEs (reference group). We also inspected the data for the presence of any brain-PAD outliers (here defined as at least +/- 3.00 SD away from the total sample mean), and subsequently excluded them (n=2) in sensitivity analyses. Post-hoc sensitivity analyses included further adjustment of the effect of PE for each of the risk factors that significantly differed across groups (Table 1) and that were suggested to be associated with brain-PAD in recent studies.^28–31^

*Additional analyses*

Further, additional analyses included a multivariable linear regression model estimating the effect of each PE sub-group, in which PE status was specified as categorical nominal predictor (i.e, three dummy variables representing ‘suspected’ ‘definite, non-clinical’, ‘definite, clinical’; where those without PEs are set as the reference group). A similar approach was used to explore the association between transient or recurring PE and brain-PAD (i.e. one dummy variable per group), where those who had no PE at neither points of assessments were the reference group. Both age and sex were added as covariates in all models. Lastly, to estimate the relative contribution of cortical and subcortical features in brain age prediction, Pearson correlation coefficients were calculated for brain-predicted and each FreeSurfer measure across the total sample or within groups.

**8. Supplementary analysis with ENIGMA-derived brain-PAD**

*PEs and brain age*

Repeating analyses with ENIGMA-derived brain-PAD as the outcome did not provide evidence for an association with PEs (Tables S3-S7 and Figure S5). Nonetheless, and regardless of statistical significance, the direction of effect (point) estimates was rather inconsistent between primary (binary/ordinal PE) and exploratory (recurring/transient PE) analyses, and observed effect sizes appeared relatively trivial (Cohen’s *d* = ± 0.04-0.09) compared to those obtained with CentileBrain-derived brain-PAD (Table S4 and Table S5). This apparent discrepancy should be interpreted with caution given the following limitations of the ENIGMA model in the current study. First, age-prediction accuracy was considerably lower that of the CentileBrain model (i.e., higher MAE/wMAE), with the female model exhibiting higher MAE than the male one. Second, systemic over-estimation of brain age across participants (regardless of sex or PE status), with no linear dependence of brain-PAD on chronological age, implying that this considerable bias was not necessarily circumvented by adding chronological age as a covariate in subsequent linear regression models for the association between PEs and brain-PAD. In addition, and regardless of lower age-prediction accuracy, it is equally important to emphasise that the ENIGMA model was trained on a primarily adult sample (18-75 years), whereas the CentileBrain was trained on a sample that covers a period from childhood through young adulthood (5-40 years). Given that adolescence is characterised by overlapping as well as unique age-related structural brain patterns relative to mid- and late- adulthood,^32–35^ the ENIGMA model may therefore carry less relevant information for accurately estimating “brain age” in the current sample of late adolescents (19-21 years).

*Relative feature importance in brain age prediction*

Nearly all FreeSurfer features were negatively correlated to ENIGMA-predicted brain age at varying degrees across the total sample (overall *r* range: -0.41 to +0.01), except from mean lateral ventricle volume which was positively correlated (r = +0.29; Table S9). On average, cortical thickness features showed relatively stronger negative correlations (mean *r* [SD] = -0.29 [0.15]) than surface area features (-0.21 [0.05]). Subcortical grey matter volume features showed a similar magnitude of negative correlations (mean *r* [SD] = -0.36 [0.11]), in addition to ICV (r = -0.33). With respect to individual brain regions, the top correlations in terms of magnitude were located bilaterally in thalamic volume, thickness of posterior cingulate, superior frontal, paracentral, precentral, and precuneal cortices, and nucleus accumbens volume (*r* range: -0.44 to -0.51). The relative strength of these top correlations was fairly comparable within the control (no-PE) and PE groups (Table S9).

| **Supplementary Tables**  **Supplementary Table S1. Characteristics of participants excluded due failed image reconstruction or quality control of image processing** | | | | | |
| --- | --- | --- | --- | --- | --- |
| **PE status** | **N** | **Mean age (SD)** | **Female, n (%)** | **Reason for exclusion, n (%)** | **PE sub-type, n(%)** |
| No PEs | 11 | 20.26 (0.62) | 4 (36.4) | IR: 3 (27.3), QC: 8 (72.7) | - |
| PEs | 9 | 20.12 (0.61) | 5 (55.6) | IR: 3 (33.3), QC: 6 (66.7) | S: 3 (33.3), D: 2 (22.2), Dc: 4 (44.5) |
| Notes:  PE: Psychotic experiences; IR: Failed image reconstruction; QC: failed quality control of cortical parcellation and/or subcortical segmentation; S: suspected PE; D: definite PE; Dc: Definite, clinical PE | | | | | |

| **Supplementary Table 2. Model performance metrics in the current sample** | | | | | | |
| --- | --- | --- | --- | --- | --- | --- |
| **Metric /**  **per model ^a^** | **Total** | | **Without PE** | | **With PE** | |
|  | **M (N=77)** | **F (N=155)** | **M (N=42)** | **F (N=73)** | **M (N=35)** | **F (N=82)** |
| ***ENIGMA model*** | | | | | | |
| **MAE (SE)** | 5.43 (0.48) * | 6.83 (0.38) * | 5.30 (0.60) * | 6.46 (0.42) * | 5.59 (0.77) * | 7.17 (0.61) * |
| **SD_AE_** | 4.18 | 4.70 | 3.91 | 3.59 | 4.53 | 5.50 |
| **RSD_AE_ (%) ^b^** | 76.98 | 68.81 | 73.77 | 55.58 | 81.04 | 76.71 |
| **wMAE ^c^** | 0.095 | 0.12 | 0.093 | 0.11 | 0.098 | 0.13 |
| **r ^d^** | 0.11 | 0.06 | 0.24 | -0.03 | -0.04 | 0.16 |
| **R^2^** | 0.012 | 0.004 | 0.06 | 0.0009 | 0.002 | 0.01 |
| ***CentileBrain mode****l* | | | | | | |
| **MAE (SE)** | 2.36 (0.19) ** | 2.47 (0.15) ** | 2.13 (0.26) ** | 2.21 (0.20)** | 2.65 (0.27)** | 2.70 (0.22)** |
| **SD_AE_** | 1.65 | 1.86 | 1.68 | 1.72 | 1.60 | 1.95 |
| **RSD_AE_ (%) ^b^** | 69.92 | 75.30 | 78.87 | 77.81 | 60.38 | 72.22 |
| **wMAE ^c^** | 0.067 | 0.071 | 0.061 | 0.063 | 0.076 | 0.077 |
| **r ^~~d~~^** | 0.13 | 0.006 | 0.07 | 0.12 | 0.12 | -0.07 |
| **R^2^** | 0.017 | 0.00003 | 0.005 | 0.014 | 0.014 | 0.005 |
| **Notes:**  ^a^ Lower MAE typically indicates better model fit, while higher *r* and *R^2^* indicates better model fit. Comparison of these metrics between brain age models is not straightforward, and they should be considered in context with underlying dataset attributes, such as the age range of training and testing set(s).^14^  ^b^ RSD_AE_ = SD_AE_ x 100 / MAE (%). This is not a formal performance metric but rather a standardised measure of how absolute errors are spread relative to the sample mean for evaluating prediction error variance.  ^c^ wMAE: MAE divided by age range of the respective training set (ENIGMA: 57 [75-18]; CentileBrain: 35 [40-5])  ^d^ Low *r* and *R^2^* for both brain-age models at least partly reflect low variance in chronological age (due to very narrow age of the testing sample), which inevitably decreases covariance between predicted and actual age.^14^  * Post-hoc t-test: MAE differed between males and females across the total sample (p=0.022), and at least to some extent among those without PE (p=0.12) or with PE (p=0.12).  ** Posthoc t-test: MAE appeared similar between males and females in the total sample (p=0.67), or among those without PE (p=0.81) or with PE (p=0.88).  MAE: mean absolute error between predicted brain age and chronological age; SD_AE_: standard deviation of absolute errors; RSD: Relative SD_AE_; r: Pearson’s correlation coefficient between brain-predicted age and chronological age; SE: standard error. R2: age variance explained by brain-predicted age. | | | | | | |

[Proceed to next page]

| **Supplementary Table S3.** Primary analyses of the association between binary or ordinal PE classification and brain-PAD, before and after exclusion of brain-PAD outliers. | | | | | | | | | | | | |
| --- | --- | --- | --- | --- | --- | --- | --- | --- | --- | --- | --- | --- |
| **Predictor** | **Outcome ^a^** | | | | | | | | | | | |
| **PE classification** | ***CentileBrain-derived brain-PAD*** | | | | | | ***ENIGMA-derived brain-PAD*** | | | | | |
| - | ***b*** | ***b* SE** | ***b* 95% CI** | **p** | ***d / R^2^ ^b^*** | ***d 95% CI ^b^*** | **b** | **bSE** | **b 95%CI** | **p** | ***d / R ^2^ ^b^*** | ***d* 95% CI *^b^*** |
| *Before exclusion of outliers ^c^* | | | | | | | *Before exclusion of outliers ^c^* | | | | | |
| Binary PE | - .62 | .39 | -1.37, .15 | .11 | -.21 | -.47, .05 | .52 | .84 | -1.13, 2.17 | .53 | .08 | - .17, .34 |
| Ordinal PE | - .26 | .18 | -.61, .09 | .15 | .009 | - | .12 | .39 | -.64, .89 | .75 | .0004 | - |
| *After exclusion of outliers (n=2) ^c^* | | | | | | | *After exclusion of outliers (n=0) ^c^* | | | | | |
| Binary PE | - .60 | .37 | -1.34, 0.14 | .11 | -.21 | -.47, .05 | - | - |  | - | - | - |
| Ordinal PE | - .28 | .17 | -.62, .06 | .11 | .011 | . - | - | - |  | - | - | - |
| Notes:  ^a^ Multiple linear regression was performed with either CentileBrain-derived brain-PAD (left) *or* ENIGMA-derived brain-PAD (right) as a continuous outcome. PE were specified as either a binary (i.e. PE versus none) *or* ordinal predictor (none > suspected > definite > definite, clinical). Effect estimates were adjusted for age and sex.  *^b^* Cohen’s d and partial R2 are reported for binary- and ordinal- PEs, respectively. 95% is reported for Cohen’s d only.  ^c^ N_total_ = 232 (n_no-PE_ = 115; n_suspected_ = 41, n_definite_ = 46; n_definite, clinical_ = 30). After outlier exclusion: N_total_ = 230 (n_no-PE_ = 114; n_suspected_ = 41, n_definite_ = 46; n_definite, clinical_ = 29). No outliers detected for ENIGMA-derived brain-PAD.  PE: Psychotic experiences; Brain-PAD: difference between brain-predicted age and chronological age (years); *b*: unstandardised regression coefficient; *SE*: standard error of *b*; *p*: p-value; 95% CI: 95% confidence interval; *d*: Cohen’s *d*; *R^2^:* proportion of variance in brain-PAD accounted by ordinal PEs. | | | | | | | | | | | | |

| **Supplementary Table S4.** Post-hoc subgroup analyses of difference in mean-brain-PAD between each PE category and those without PE, before- and after- exclusion of brain-PAD outliers | | | | | | | | | | | | |
| --- | --- | --- | --- | --- | --- | --- | --- | --- | --- | --- | --- | --- |
| **Predictor** | **Outcome ^a^** | | | | | | | | | | | |
| **(PE sub-group)** | ***CentileBrain-derived brain-PAD*** | | | | | | ***ENIGMA-derived brain-PAD*** | | | | | |
| - | **b** | **SE** | **b 95% CI** | **p** | ***d*** | ***d 95% CI*** | **b** | **SE** | **b 95%CI** | **p** | ***d*** | ***d 95% CI*** |
| *Before exclusion of outliers ^b^* | | | | | | | *Before exclusion of outliers ^b^* | | | | | |
| Suspected | - .46 | .55 | -1.53, .62 | .40 | - .13 | -.48, .23 | 1.00 | 1.18 | -1.33, 3.32 | .40 | .13 | -.23, .48 |
| Definite | - .82 | .53 | -1.85, .22 | .12 | - .23 | -.57, .11 | .23 | 1.14 | -2.01, 2.47 | .84 | .03 | -.31, .37 |
| Definite, Clinical | - .53 | .61 | -1.74, .67 | .39 | - .14 | -54, .26 | .28 | 1.33 | -2.33, 2.89 | .83 | .03 | -.37, .43 |
| *After exclusion of outliers (n=2) ^c^* | | | | | | | *After exclusion of outliers (n=0) ^c^* | | | | | |
| Suspected | - .41 | .53 | -1.46, .63 | .43 | - .12 | -.47, .23 | - | - | - | - | - | - |
| Definite | - .70 | .51 | -1.70, 0.30 | .17 | - .20 | -.56, .14 | - | - | - | - | - | - |
| Definite, clinical | - .72 | .60 | -1.90, .46 | .23 | - .20 | -.61, .21 | - | - | - | - | - | - |
| Notes:  ^a^ Multivariable linear regression was performed with either CentileBrain-derived brain-PAD (left; primary) or ENIGMA-derived brain-PAD (right) as outcome. Suspected, definite, and definite, clinical PEs were specified as a categorical nominal predictor (i.e., 3 dummy predictors) and those without PE were set as the reference group. Effect estimates were adjusted for age and sex.  ^b^ N_total_ = 232 (n_suspected_ = 41, n_defite, non-clinical_ = 46; n_defite, clinical_ = 30; n_no-PE_ = 115)  ^c^ N_total_ = 230 (n_suspected_ = 41, n_defite, non-clinical_ = 46; n_defite, clinical_ = 29; n_no-PE_ = 114). No outliers were identified for ENIGMA-derived brain-PAD.  PE: Psychotic experiences; Brain-PAD: difference between brain-predicted age and chronological age (years); *b*: unstandardised regression coefficient;  *SE*: standard error of *b*; *p*: p-value; 95% CI: 95% confidence interval; *d*: Cohen’s *d* | | | | | | | | | | | | |

| **Supplementary Table S5**. Exploratory analyses of the association between transient or recurring PE and brain-PAD, before and after exclusion of brain-PAD outliers. | | | | | | | | | | | | |
| --- | --- | --- | --- | --- | --- | --- | --- | --- | --- | --- | --- | --- |
| **Predictor** | **Outcome ^a^** | | | | | | | | | | | |
|  | ***CentileBrain-derived brain-PAD*** | | | | | | ***ENIGMA-derived brain-PAD*** | | | | | |
| - | **b** | **SE** | **b 95% CI** | **p** | ***d*** | ***d 95% CI*** | **b** | **bSE** | **b 95% CI** | **p** | ***d*** | ***d 95% CI*** |
| *Before exclusion of outliers ^b^* | | | | | | | *Before exclusion of outliers ^b^* | | | | | |
| Transient PE | - .56 | .44 | -1.44, 0.30 | .20 | -.18 | -.49, .12 | -.61 | .98 | -2.53, 1.32 | .54 | - .09 | -.40, .22 |
| Recurring PE | - .61 | .56 | -1.70, 0.48 | .27 | -.18 | -.56, .21 | -.28 | .23 | -2.70, 2.14 | .82 | - .04 | -.42, .35 |
| *After exclusion of outliers (n=1) ^c^* | | | | | | | *After exclusion of outliers (n=0) ^c^* | | | | | |
| Transient PE | - .46 | .43 | -1.32, 0.39 | .29 | -.15 | -.46, .16 | - | - | - | - | - | - |
| Recurring PE | - .51 | .54 | -1.58, 0.57 | .36 | -.15 | -.53, .24 | - | - | - | - | - | - |
| Notes:  ^a^ Multivariable linear regression was performed with either CentileBrain-derived brain-PAD (left; primary) or ENIGMA-derived brain-PAD (right) as outcome. Transient and recurring PEs were specified as categorical nominal variables (i.e. 2 dummy predictors) with those without PE at both points of ascertainment set as the reference group. Effect estimates were adjusted for age and sex.  ^b^ N_total_ = 202 (n_transient_= 73, n_reccuring_ = 36; n_no-PE_ = 93)  ^c^ N_total_ = 201 (n_transient_= 73, n_reccuring_ = 36; n_no-PE_ = 92). No outliers were identified for ENIGMA-derived brain-PAD  PE: Psychotic experiences; Brain-PAD: difference between brain-predicted age and chronological age (years); *b*: unstandardised regression coefficient;  *SE*: standard error of *b*; *p*: p-value; 95% CI: 95% confidence interval; *d*: Cohen’s *d* | | | | | | | | | | | | |

| **Supplementary Table S6**. Sensitivity analyses of the association between brain-PAD and PEs not attributable to sleep/fever, before- and after- exclusion of brain-PAD outliers. | | | | | | | | | | | | |
| --- | --- | --- | --- | --- | --- | --- | --- | --- | --- | --- | --- | --- |
| **Predictor** | **Outcome ^a^** | | | | | | | | | | | |
|  | ***CentileBrain-derived brain-PAD*** | | | | | | ***ENIGMA-derived brain-PAD*** | | | | | |
| - | **b** | **b SE** | **b 95% CI** | **p** | ***d / R^2^ ^d^*** | ***d* 95% CI *^d^*** | **b** | **b SE** | **b 95% CI** | **p** | ***d/R^2^*** | ***d 95% CI*** |
| *Before exclusion of outliers ^b^* | | | | | | | *Before exclusion of outliers ^b^* | | | | | |
| PE vs. no-PE | - .45 | .39 | -1.22, 0.32 | .26 | - .16 | -.41, .11 | .55 | .84 | -1.11, 2.21 | .52 | .09 | -.17, .34 |
| Ordinal PE | - .21 | .18 | -0.57, 0.14 | .24 | .006 | - | .09 | .39 | -0.67, 0.85 | .81 | .0002 | - |
| *After exclusion of outliers (n=2) ^c^* | | | | | | | *After exclusion of outliers (n=0) ^c^* | | | | | |
| PE vs. no-PE | - .45 | .38 | -1.19, 0.29 | .23 | - .16 | -.42, .10 | - | - | - | - | - | - |
| Ordinal PE | - .25 | .17 | -0.59, 0.10 | .16 | .009 | - | - | - | - | - | - | - |
| Notes:  ^a^ Multivariable linear regression was performed with either CentileBrain-derived brain-PAD (left; primary) or ENIGMA-derived brain-PAD (right; sensitivity) as outcome. PEs were specified as either a binary (i.e. PE vs. no-PE) or ordinal predictor (none > suspected > definite, non-clinical > definite, clinical) predictor. Effect estimates were adjusted for age and sex.  ^b^ N_total_ = 232; n_suspected_ = 38, n_definite, non-clinical_ = 33; n_defite, clinical_ = 30; n_no-PE_ = 131. Those with PEs solely attributed to sleep or fever were reclassified to no-PE (controls).  ^c^ N_total_ = 232; n_suspected_ = 38, n_definite, non-clinical_ = 33; n_defite, clinical_ = 29; n_no-PE_ = 130. No outliers were identified for ENIGMA-derived brain-PAD.  ^d^ Cohen’s d and partial R2 are reported for binary- and ordinal- PEs, respectively. 95% is reported for Cohen’s d only.  PE: Psychotic experiences; Brain-PAD: difference between brain-predicted age and chronological age (years); *b*: unstandardised regression coefficient; b*SE*: standard error of *b*; *p*: p-value; *d*: Cohen’s *d*; 95% CI: 95% confidence interval of b or d; *R^2^* proportion of variance in brain-PAD accounted by PEs (i.e. partial R2). | | | | | | | | | | | | |

| **Supplementary Table S7. Sensitivity analyses of association between PEs on brain-PAD additionally adjusting for potential confounders (risk-factors)** | | | | | |
| --- | --- | --- | --- | --- | --- |
| **Covariate** | **Main predictor** | **Outcome ^a^** | | | |
| **Risk factor** | **Binary/ordinal PE** | **CentileBrain-derived-brain-PAD** | | **ENIGMA-derived brain-PAD** | |
| **-** | **-** | **b_PE_** | **b_risk-factor_** | **b_PE_** | **b_risk-factor_** |
| Birth weight (g) ^b^ | Binary | -0.85 ± 0.40 (**0.038)** | -0.0001 ± 0.0004(0.77) | 0.75 ± 0.89 (0.40) | 0.0001 ± 0.0009 (0.90) |
|  | Ordinal | -0.35 ± 0.19 (0.066) | -0.0001 ± 0.0004(0.74) | 0.22 ± 0.42 (0.60) | 8x10^-05^ ± 9x10^-4^ (0.93) |
| Maternal education ^c^ | Binary | -0.60 ± 0.41 (0.16) | 0.37 ± 0.27 (0.16) | 1.03 ± 0.89 (0.25) | 0.28 ± 0.35 (0.43) |
|  | Ordinal | -0.27 ± 0.19 (0.15) | 0.39 ± 0.26 (0.15) | 0.29 ± 0.41 (0.48) | 0.20 ± 0.58 (0.73) |
| Parental social class ^d^ | Binary | -0.88 ± 0.40 (**0.031**) | -0.85 ± 0.61 (0.16) | 0.60 ± 0.90 (0.51) | -1.64 ± 1.35 (0.23) |
|  | Ordinal | -0.43 ± 0.19 (**0.024**) | -0.94 ± 0.61 (0.13) | 0.15 ± 0.43 (0.73) | -1.68 ± 1.37 (0.22) |
| Childhood IQ ^e^ | Binary | -0.67 ± 0.40 (0.096) | 0.02 ± 0.01 (0.24) | 0.36 ± .89 (0.68) | 0.03 ± 0.03 (0.22) |
|  | Ordinal | -0.35 ± 0.19 (0.066) | 0.01 ± 0.01 (0.26) | 0.0009 ± 0.42 (1.0) | 0.03 ± 0.03 (0.25) |
| Statistics presented: unstandardised coefficient (b) ± standard error (p-value for null hypothesis testing) from a multivariable linear regression for the main effect of PE (binary or ordinal) adjusted for an additional covariate (PE risk-factor) on brain-PAD, in addition to age and sex. b_risk-factor_ represents the main effect of the additional covariate/risk-factor. Bold p-values indicate significance at α = .05.  ^a^ Multivariable regression was performed with either CentileBrain-derived brain-PAD (right) or ENIGMA-derived brain-PAD (left) as outcome.PE were specified as either a binary (i.e. PE versus none; model 1) *or* ordinal predictor (none > suspected > definite > definite, clinical; model 2).  ^b^ Birth weight as specified as continuous predictor (grams). Analytical sample size was N=218 (none = 107; suspected PE = 39; definite PE = 44; definite, clinical = 29; total PE = 112).  ^c^ Maternal education was specified as a categorical ordinal predictor (None/CSE/vocational > O-levels > A-levels/degree). Analytical size sample was N=217 (none = 107; suspected PE = 37; definite PE = 45; definite, clinical = 28; total PE = 110).  ^d^ Parental social class was specified as binary predictor (I/II/III-non manual vs. III-manual/IV/V as reference). Analytical sample size was N=206 (none = 102; suspected PE = 36; definite PE= 44, definite, clinical = 24; total N= 104).  ^e^ Childhood IQ was specified as a continuous predictor (full WISC-III scale). Analytical sample size was N=201 (none = 104; suspected PE = 33; definite PE = 42, definite, clinical = 22; total PE= 97). | | | | | |

| **Supplementary Table S8. Correlation between cortical and subcortical features and CentileBrain-predicted age in the current sample** | | | | |
| --- | --- | --- | --- | --- |
| **No.** | **Freesurfer feature** | ***r _T_*_otal_** | ***r* _No-PE_** | **r _PE_** |
| 1 | L_precuneus_thickavg | -0.41 | -0.37 | -0.45 |
| 2 | R_posteriorcingulate_thickavg | -0.41 | -0.31 | -0.49 |
| 3 | R_superiorfrontal_thickavg | -0.38 | -0.27 | -0.47 |
| 4 | L_inferiorparietal_thickavg | -0.38 | -0.3 | -0.45 |
| 5 | R_precuneus_thickavg | -0.37 | -0.22 | -0.48 |
| 6 | R_superiorparietal_thickavg | -0.36 | -0.29 | -0.39 |
| 7 | L_superiorparietal_thickavg | -0.34 | -0.33 | -0.35 |
| 8 | L_superiorfrontal_thickavg | -0.33 | -0.33 | -0.33 |
| 9 | L_posteriorcingulate_thickavg | -0.32 | -0.21 | -0.39 |
| 10 | R_paracentral_thickavg | -0.32 | -0.25 | -0.37 |
| 11 | R_rostralmiddlefrontal_thickavg | -0.31 | -0.32 | -0.29 |
| 12 | L_rostralmiddlefrontal_thickavg | -0.29 | -0.31 | -0.26 |
| 13 | R_inferiorparietal_thickavg | -0.28 | -0.22 | -0.32 |
| 14 | L_bankssts_thickavg | -0.28 | -0.22 | -0.33 |
| 15 | L_parsopercularis_thickavg | -0.27 | -0.3 | -0.25 |
| 16 | R_insula_thickavg | -0.27 | -0.2 | -0.33 |
| 17 | L_parsopercularis_surfavg | -0.26 | -0.14 | -0.35 |
| 18 | R_bankssts_thickavg | -0.26 | -0.04 | -0.38 |
| 19 | L_superiortemporal_thickavg | -0.26 | -0.15 | -0.35 |
| 20 | R_superiortemporal_thickavg | -0.25 | -0.02 | -0.42 |
| 21 | R_caudalmiddlefrontal_thickavg | -0.25 | -0.13 | -0.34 |
| 22 | R_caudalanteriorcingulate_thickavg | -0.25 | -0.31 | -0.18 |
| 23 | R_cuneus_thickavg | -0.24 | -0.11 | -0.35 |
| 24 | Rcaud | -0.24 | -0.12 | -0.34 |
| 25 | L_paracentral_thickavg | -0.24 | -0.22 | -0.25 |
| 26 | L_supramargil_thickavg | -0.23 | -0.19 | -0.26 |
| 27 | R_inferiorparietal_surfavg | -0.23 | -0.17 | -0.29 |
| 28 | L_lateraloccipital_thickavg | -0.23 | -0.16 | -0.3 |
| 29 | L_cuneus_thickavg | -0.23 | -0.26 | -0.22 |
| 30 | R_bankssts_surfavg | -0.23 | -0.11 | -0.33 |
| 31 | R_middletemporal_thickavg | -0.23 | -0.11 | -0.32 |
| 32 | R_supramargil_thickavg | -0.23 | -0.19 | -0.24 |
| 33 | L_caudalmiddlefrontal_thickavg | -0.22 | -0.13 | -0.31 |
| 34 | R_lateralorbitofrontal_thickavg | -0.22 | -0.25 | -0.22 |
| 35 | L_postcentral_surfavg | -0.22 | -0.11 | -0.32 |
| 36 | L_frontalpole_thickavg | -0.22 | -0.16 | -0.27 |
| 37 | L_parstriangularis_thickavg | -0.22 | -0.26 | -0.19 |
| 38 | R_parsopercularis_thickavg | -0.22 | -0.16 | -0.27 |
| 39 | L_supramargil_surfavg | -0.21 | -0.14 | -0.32 |
| 40 | R_postcentral_thickavg | -0.21 | -0.17 | -0.23 |
| 41 | L_paracentral_surfavg | -0.21 | -0.15 | -0.25 |
| 42 | R_postcentral_surfavg | -0.21 | -0.11 | -0.32 |
| 43 | R_frontalpole_thickavg | -0.21 | -0.3 | -0.12 |
| 44 | R_parsorbitalis_thickavg | -0.21 | -0.15 | -0.25 |
| 45 | L_isthmuscingulate_surfavg | -0.2 | -0.13 | -0.31 |
| 46 | R_middletemporal_surfavg | -0.2 | -0.17 | -0.24 |
| 47 | R_parstriangularis_thickavg | -0.2 | -0.15 | -0.24 |
| 48 | Lpal | -0.2 | -0.02 | -0.33 |
| 49 | L_middletemporal_thickavg | -0.2 | -0.19 | -0.19 |
| 50 | L_parsorbitalis_thickavg | -0.19 | -0.2 | -0.19 |
| 51 | Rput | -0.19 | -0.01 | -0.32 |
| 52 | R_superiorparietal_surfavg | -0.19 | -0.12 | -0.23 |
| 53 | L_posteriorcingulate_surfavg | -0.18 | -0.11 | -0.25 |
| 54 | R_superiortemporal_surfavg | -0.18 | -0.04 | -0.31 |
| 55 | R_transversetemporal_thickavg | -0.18 | -0.11 | -0.22 |
| 56 | L_lateralorbitofrontal_thickavg | -0.18 | -0.14 | -0.21 |
| 57 | L_middletemporal_surfavg | -0.18 | -0.11 | -0.25 |
| 58 | L_parstriangularis_surfavg | -0.18 | -0.14 | -0.22 |
| 59 | Lcaud | -0.18 | -0.03 | -0.29 |
| 60 | L_bankssts_surfavg | -0.18 | -0.19 | -0.16 |
| 61 | L_inferiortemporal_surfavg | -0.17 | -0.09 | -0.25 |
| 62 | L_insula_thickavg | -0.17 | -0.07 | -0.27 |
| 63 | R_parahippocampal_surfavg | -0.17 | -0.26 | -0.09 |
| 64 | L_superiorparietal_surfavg | -0.17 | -0.12 | -0.23 |
| 65 | L_precentral_surfavg | -0.17 | 0.01 | -0.3 |
| 66 | R_precuneus_surfavg | -0.17 | -0.11 | -0.22 |
| 67 | R_isthmuscingulate_surfavg | -0.16 | -0.07 | -0.28 |
| 68 | R_precentral_thickavg | -0.16 | -0.17 | -0.17 |
| 69 | L_postcentral_thickavg | -0.16 | -0.13 | -0.19 |
| 70 | R_posteriorcingulate_surfavg | -0.16 | -0.14 | -0.21 |
| 71 | R_precentral_surfavg | -0.16 | -0.04 | -0.26 |
| 72 | R_lateraloccipital_thickavg | -0.16 | -0.1 | -0.2 |
| 73 | Lput | -0.16 | 0.05 | -0.33 |
| 74 | R_supramargil_surfavg | -0.16 | -0.1 | -0.23 |
| 75 | L_precentral_thickavg | -0.16 | -0.16 | -0.16 |
| 76 | L_superiorfrontal_surfavg | -0.16 | -0.02 | -0.29 |
| 77 | R_frontalpole_surfavg | -0.15 | -0.09 | -0.22 |
| 78 | R_parsopercularis_surfavg | -0.15 | -0.05 | -0.24 |
| 79 | L_superiortemporal_surfavg | -0.15 | -0.08 | -0.24 |
| 80 | R_transversetemporal_surfavg | -0.15 | -0.08 | -0.23 |
| 81 | R_inferiortemporal_surfavg | -0.15 | -0.08 | -0.25 |
| 82 | L_rostralmiddlefrontal_surfavg | -0.15 | -0.11 | -0.22 |
| 83 | L_precuneus_surfavg | -0.15 | -0.15 | -0.18 |
| 84 | L_inferiortemporal_thickavg | -0.15 | -0.2 | -0.1 |
| 85 | L_transversetemporal_surfavg | -0.15 | -0.06 | -0.24 |
| 86 | R_inferiortemporal_thickavg | -0.15 | -0.18 | -0.13 |
| 87 | Rthal | -0.15 | 0.02 | -0.3 |
| 88 | R_rostralmiddlefrontal_surfavg | -0.15 | -0.07 | -0.24 |
| 89 | L_fusiform_surfavg | -0.14 | -0.02 | -0.24 |
| 90 | L_rostralanteriorcingulate_thickavg | -0.14 | -0.27 | -0.05 |
| 91 | L_lingual_surfavg | -0.13 | -0.12 | -0.17 |
| 92 | R_lateralorbitofrontal_surfavg | -0.13 | -0.06 | -0.22 |
| 93 | L_inferiorparietal_surfavg | -0.13 | -0.1 | -0.19 |
| 94 | L_caudalanteriorcingulate_thickavg | -0.13 | -0.06 | -0.18 |
| 95 | R_paracentral_surfavg | -0.13 | -0.05 | -0.21 |
| 96 | R_caudalmiddlefrontal_surfavg | -0.13 | 0.01 | -0.22 |
| 97 | Raccumb | -0.13 | 0.06 | -0.29 |
| 98 | Laccumb | -0.12 | -0.07 | -0.19 |
| 99 | R_medialorbitofrontal_thickavg | -0.12 | -0.12 | -0.11 |
| 100 | Lthal | -0.12 | 0.03 | -0.26 |
| 101 | Rpal | -0.12 | 0.05 | -0.25 |
| 102 | R_rostralanteriorcingulate_thickavg | -0.12 | -0.15 | -0.09 |
| 103 | R_medialorbitofrontal_surfavg | -0.11 | -0.03 | -0.21 |
| 104 | R_parsorbitalis_surfavg | -0.11 | -0.06 | -0.2 |
| 105 | L_parahippocampal_surfavg | -0.1 | -0.12 | -0.08 |
| 106 | L_parsorbitalis_surfavg | -0.1 | -0.02 | -0.2 |
| 107 | R_parstriangularis_surfavg | -0.09 | 0.06 | -0.24 |
| 108 | L_lateralorbitofrontal_surfavg | -0.09 | -0.05 | -0.17 |
| 109 | R_superiorfrontal_surfavg | -0.09 | 0.06 | -0.23 |
| 110 | R_caudalanteriorcingulate_surfavg | -0.09 | -0.08 | -0.12 |
| 111 | L_rostralanteriorcingulate_surfavg | -0.08 | 0.03 | -0.18 |
| 112 | L_caudalmiddlefrontal_surfavg | -0.08 | 0.11 | -0.27 |
| 113 | R_pericalcarine_thickavg | -0.08 | 0.07 | -0.2 |
| 114 | L_pericalcarine_thickavg | -0.08 | 0.02 | -0.15 |
| 115 | R_fusiform_surfavg | -0.08 | 0.11 | -0.25 |
| 116 | Ramyg | -0.08 | 0.01 | -0.16 |
| 117 | R_isthmuscingulate_thickavg | -0.07 | -0.07 | -0.05 |
| 118 | R_lingual_surfavg | -0.07 | -0.06 | -0.09 |
| 119 | L_insula_surfavg | -0.07 | 0.06 | -0.18 |
| 120 | L_medialorbitofrontal_thickavg | -0.06 | -0.03 | -0.08 |
| 121 | L_caudalanteriorcingulate_surfavg | -0.05 | 0.03 | -0.1 |
| 122 | Lamyg | -0.05 | 0.02 | -0.13 |
| 123 | R_cuneus_surfavg | -0.05 | 0 | -0.14 |
| 124 | L_lingual_thickavg | -0.04 | 0.03 | -0.1 |
| 125 | R_lateraloccipital_surfavg | -0.04 | -0.03 | -0.1 |
| 126 | L_frontalpole_surfavg | -0.04 | -0.02 | -0.08 |
| 127 | L_medialorbitofrontal_surfavg | -0.04 | -0.01 | -0.1 |
| 128 | R_lingual_thickavg | -0.04 | 0.02 | -0.07 |
| 129 | L_lateraloccipital_surfavg | -0.02 | 0.07 | -0.11 |
| 130 | L_pericalcarine_surfavg | -0.02 | 0 | -0.07 |
| 131 | L_transversetemporal_thickavg | -0.02 | 0.11 | -0.09 |
| 132 | L_isthmuscingulate_thickavg | -0.01 | -0.03 | 0.01 |
| 133 | R_rostralanteriorcingulate_surfavg | -0.01 | 0 | -0.06 |
| 134 | R_temporalpole_thickavg | 0 | 0.06 | -0.07 |
| 135 | R_entorhil_surfavg | 0 | 0.11 | -0.11 |
| 136 | L_parahippocampal_thickavg | 0 | -0.03 | 0.02 |
| 137 | L_temporalpole_surfavg | 0 | 0.02 | -0.02 |
| 138 | R_pericalcarine_surfavg | 0 | -0.01 | -0.01 |
| 139 | R_temporalpole_surfavg | 0.02 | 0.06 | -0.05 |
| 140 | R_insula_surfavg | 0.03 | 0.16 | -0.09 |
| 141 | L_entorhil_surfavg | 0.04 | 0.07 | -0.03 |
| 142 | R_fusiform_thickavg | 0.04 | 0.12 | -0.01 |
| 143 | L_entorhil_thickavg | 0.06 | 0.05 | 0.06 |
| 144 | L_cuneus_surfavg | 0.06 | 0.16 | -0.06 |
| 145 | L_fusiform_thickavg | 0.08 | 0.02 | 0.11 |
| 146 | Lhippo | 0.08 | 0.2 | -0.07 |
| 147 | L_temporalpole_thickavg | 0.12 | 0.05 | 0.19 |
| 148 | R_parahippocampal_thickavg | 0.13 | 0.13 | 0.14 |
| 149 | Rhippo | 0.14 | 0.21 | 0.07 |
| 150 | R_entorhil_thickavg | 0.15 | 0.15 | 0.15 |
| **Notes/Abbreviations**  Correlation coefficients are listed in the order of magnitude across the total sample (r_Total_), ranging from moderately negative (top) to weakly positive(bottom). Note that the CentileBrain model includes a separate FreeSurfer region-of-interest for each hemisphere, therefore these correlations are presented as such (with prefix R or L for right and left hemisphere respectively).  r_Total:_ correlation between brain-predicted age and each FreeSurfer ROI across the total sample (N=232);  *r*_No-PE:_ correlation between brain-predicted age and each FreeSurfer ROI among those without PE (N=115);  *r*_PE:_ correlation between brain-predicted age and each FreeSurfer ROI among those with PE (N=117). | | | | |

| **Supplementary Table S9. Correlation between cortical and subcortical features and ENIGMA -predicted age in the current sample** | | | | |
| --- | --- | --- | --- | --- |
| **No.** | **Freesurfer feature** | ***r_t_*_otal_** | ***r*_No-PE_** | **r_PE_** |
| 1 | Mthal | -0.51 | -0.43 | -0.58 |
| 2 | M_posteriorcingulate_thickavg | -0.51 | -0.48 | -0.53 |
| 3 | M_superiorfrontal_thickavg | -0.50 | -0.51 | -0.51 |
| 4 | M_paracentral_thickavg | -0.48 | -0.45 | -0.50 |
| 5 | M_precentral_thickavg | -0.47 | -0.47 | -0.47 |
| 6 | M_precuneus_thickavg | -0.46 | -0.36 | -0.54 |
| 7 | Maccumb | -0.44 | -0.39 | -0.47 |
| 8 | M_inferiorparietal_thickavg | -0.43 | -0.38 | -0.48 |
| 9 | M_parsopercularis_thickavg | -0.42 | -0.42 | -0.43 |
| 10 | Mcaud | -0.41 | -0.32 | -0.47 |
| 11 | M_cuneus_thickavg | -0.39 | -0.33 | -0.44 |
| 12 | M_lateraloccipital_thickavg | -0.39 | -0.34 | -0.44 |
| 13 | M_supramarginal_thickavg | -0.39 | -0.38 | -0.42 |
| 14 | M_superiorparietal_thickavg | -0.39 | -0.34 | -0.44 |
| 15 | M_parstriangularis_thickavg | -0.39 | -0.35 | -0.41 |
| 16 | M_caudalmiddlefrontal_thickavg | -0.37 | -0.27 | -0.46 |
| 17 | M_insula_thickavg | -0.36 | -0.43 | -0.31 |
| 18 | Mput | -0.36 | -0.32 | -0.39 |
| 19 | M_postcentral_thickavg | -0.36 | -0.33 | -0.38 |
| 20 | M_superiortemporal_thickavg | -0.35 | -0.24 | -0.42 |
| 21 | M_middletemporal_thickavg | -0.34 | -0.41 | -0.30 |
| 22 | ICV | -0.33 | -0.26 | -0.38 |
| 23 | M_parsorbitalis_thickavg | -0.32 | -0.34 | -0.30 |
| 24 | Mpal | -0.31 | -0.22 | -0.38 |
| 25 | M_rostralmiddlefrontal_thickavg | -0.31 | -0.29 | -0.34 |
| 26 | M_posteriorcingulate_surfavg | -0.28 | -0.27 | -0.28 |
| 27 | M_isthmuscingulate_thickavg | -0.28 | -0.28 | -0.28 |
| 28 | M_precentral_surfavg | -0.27 | -0.19 | -0.33 |
| 29 | M_caudalmiddlefrontal_surfavg | -0.27 | -0.14 | -0.37 |
| 30 | M_middletemporal_surfavg | -0.26 | -0.20 | -0.31 |
| 31 | M_inferiorparietal_surfavg | -0.26 | -0.21 | -0.30 |
| 32 | M_superiorfrontal_surfavg | -0.26 | -0.15 | -0.34 |
| 33 | M_transversetemporal_thickavg | -0.26 | -0.14 | -0.34 |
| 34 | M_caudalanteriorcingulate_thickavg | -0.26 | -0.29 | -0.24 |
| 35 | M_bankssts_thickavg | -0.26 | -0.17 | -0.32 |
| 36 | M_fusiform_surfavg | -0.25 | -0.22 | -0.27 |
| 37 | M_rostralmiddlefrontal_surfavg | -0.25 | -0.18 | -0.30 |
| 38 | M_isthmuscingulate_surfavg | -0.25 | -0.21 | -0.27 |
| 39 | M_parsopercularis_surfavg | -0.24 | -0.19 | -0.28 |
| 40 | Mhippo | -0.24 | -0.24 | -0.25 |
| 41 | M_inferiortemporal_surfavg | -0.24 | -0.13 | -0.32 |
| 42 | M_lingual_surfavg | -0.24 | -0.20 | -0.27 |
| 43 | M_supramarginal_surfavg | -0.24 | -0.15 | -0.31 |
| 44 | M_postcentral_surfavg | -0.24 | -0.17 | -0.28 |
| 45 | M_lateralorbitofrontal_thickavg | -0.23 | -0.29 | -0.19 |
| 46 | M_parstriangularis_surfavg | -0.23 | -0.29 | -0.19 |
| 47 | M_bankssts_surfavg | -0.23 | -0.26 | -0.22 |
| 48 | M_rostralanteriorcingulate_thickavg | -0.23 | -0.25 | -0.22 |
| 49 | M_lateralorbitofrontal_surfavg | -0.23 | -0.22 | -0.23 |
| 50 | Mamyg | -0.23 | -0.23 | -0.23 |
| 51 | M_superiorparietal_surfavg | -0.22 | -0.23 | -0.21 |
| 52 | M_parahippocampal_surfavg | -0.22 | -0.17 | -0.27 |
| 53 | M_superiortemporal_surfavg | -0.22 | -0.14 | -0.28 |
| 54 | M_pericalcarine_thickavg | -0.22 | -0.23 | -0.21 |
| 55 | M_transversetemporal_surfavg | -0.21 | -0.20 | -0.21 |
| 56 | M_precuneus_surfavg | -0.21 | -0.20 | -0.21 |
| 57 | M_medialorbitofrontal_surfavg | -0.20 | -0.11 | -0.26 |
| 58 | M_lateraloccipital_surfavg | -0.20 | -0.10 | -0.27 |
| 59 | M_parsorbitalis_surfavg | -0.20 | -0.21 | -0.18 |
| 60 | M_frontalpole_surfavg | -0.18 | -0.04 | -0.29 |
| 61 | M_rostralanteriorcingulate_surfavg | -0.18 | -0.17 | -0.18 |
| 62 | M_paracentral_surfavg | -0.17 | -0.07 | -0.24 |
| 63 | M_caudalanteriorcingulate_surfavg | -0.16 | -0.18 | -0.15 |
| 64 | M_insula_surfavg | -0.15 | -0.13 | -0.16 |
| 65 | M_inferiortemporal_thickavg | -0.15 | -0.27 | -0.06 |
| 66 | M_temporalpole_surfavg | -0.15 | -0.15 | -0.15 |
| 67 | M_pericalcarine_surfavg | -0.14 | -0.10 | -0.18 |
| 68 | M_cuneus_surfavg | -0.13 | -0.06 | -0.18 |
| 69 | M_fusiform_thickavg | -0.11 | -0.07 | -0.14 |
| 70 | M_lingual_thickavg | -0.11 | -0.19 | -0.06 |
| 71 | M_entorhinal_surfavg | -0.09 | -0.06 | -0.11 |
| 72 | M_frontalpole_thickavg | -0.06 | -0.08 | -0.05 |
| 73 | M_temporalpole_thickavg | -0.04 | 0.02 | -0.10 |
| 74 | M_entorhinal_thickavg | -0.03 | 0.00 | -0.06 |
| 75 | M_parahippocampal_thickavg | -0.02 | -0.02 | -0.01 |
| 76 | M_medialorbitofrontal_thickavg | 0.01 | -0.02 | 0.04 |
| 77 | Mvent | 0.29 | 0.29 | 0.28 |
| Notes/Abbreviations  Correlation coefficients are listed in the order of magnitude across the total sample (r_Total_), ranging from moderately negative (top) to moderate positive (bottom). Note that the ENIGMA model includes FreeSurfer region-of-interest that  were averaged across the two hemispheres (i.e. R + L / 2), therefore these correlations are presented as such (and thus the prefix “M”).  r_Total:_ correlation between brain-predicted age and each FreeSurfer ROI across the total sample (N=232);  *r*_No-PE:_ correlation between brain-predicted age and each FreeSurfer ROI among those without PE (N=115);  *r*_PE:_ correlation between brain-predicted age and each FreeSurfer ROI among those with PE (N=117).  ROI: Region of Interest | | | | |

**Supplementary Figures**

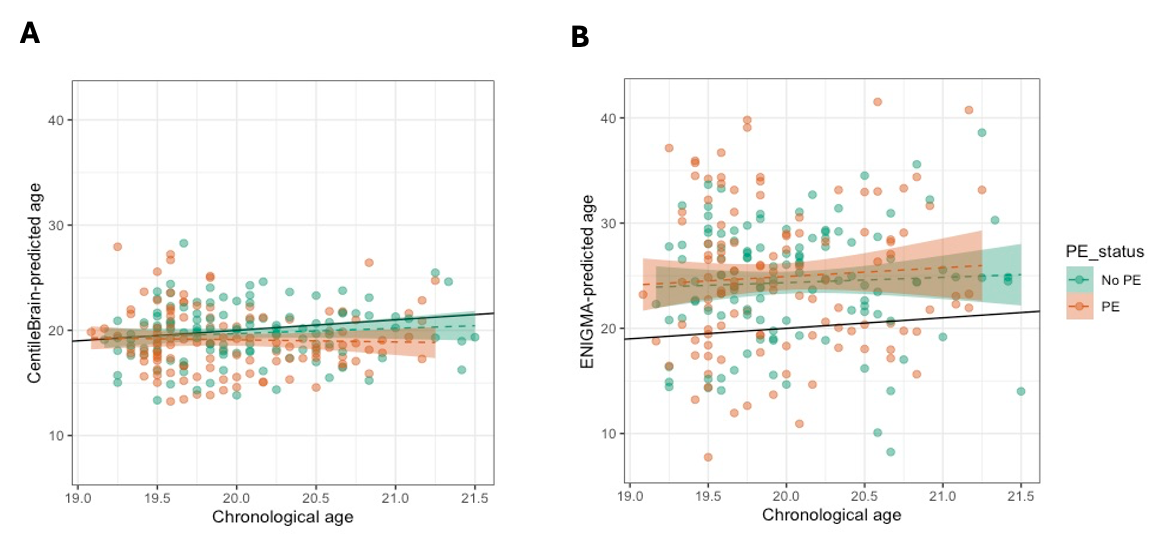

**Supplementary Figure S1.** Chronological age versus brain-predicted age with respect to psychotic experiences (PE).

**(A)** Chronological age versus brain-predicted age for the CentileBrain model. The black line represents a (hypothetical) exact linear relationship between chronological age and brain-predicted age (years). Dashed lines represent the actual fit (with standard error shading) between chronological and brain-predicted age for those with PE or without PEs. A tendency toward underestimation of brain-predicted age for relatively older participants (> ~ 20 years) was observed in those with and without PE. **(B)** Similar to (A), chronological age versus brain-predicted age for the ENIGMA model. A considerable overestimation of brain-predicted age across chronological age was observed in those with- and without PE.

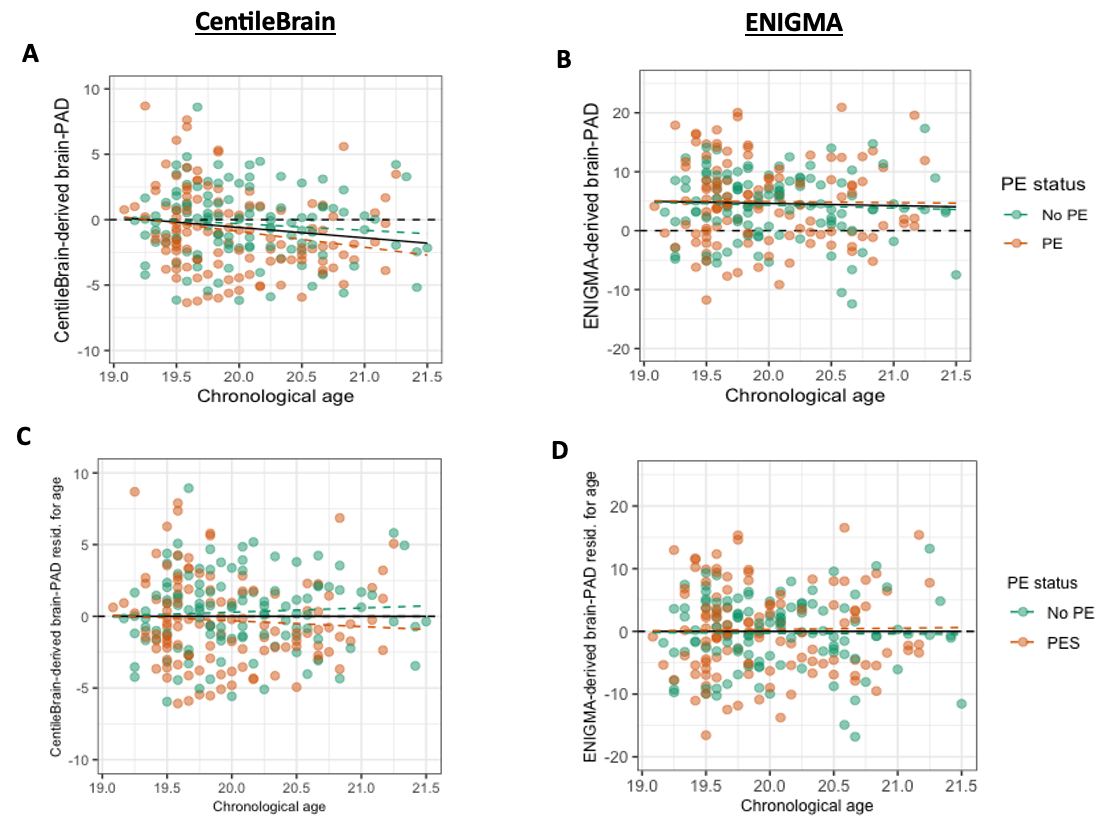

**Supplementary Figure S2. Age-related bias in brain age prediction.** Relationship between chronological age and brain-predicted age difference (brain-PAD) derived from the Centilebrain model (left) or ENIGMA model (right). A and B show brain-PAD (years) against chronological age with respect to PE status for the CentileBrain model and ENIGMA model, respectively. **(A)** The negative correlation between CentileBrain-derived brain-PAD and chronological age (r = - 0.15; p = 0.03) is due to a systematic underestimation in older individuals (> ~ 20.0 years; mean brain-PAD = -0.60 years). **(B)** While there was a lack of a linear relationship between ENIGMA-derived brain-PAD and chronological age (r = -0.03, p= 0.62), the ENIGMA model systematically overestimated brain age across the current sample (mean brain-PAD = + 4.62 years). C and D illustrate linear correction of age-bias by residualizing brain-PAD scores for chronological age**. (C)** After residualisation, CentileBrain-derived brain-PAD was no longer correlated to chronological age (r= -2x10^-16^). **(D)** After residualisation, ENIGMA-derived brain-PAD scores are centred around zero (r = 2x10^-16^). A-D: the black line shows the linear fit applied to model age-related bias across all participants, whereas linear fit within those with PE and those without PE is depicted with orange and green dashed lines, respectively. Note the difference in scale range of the y-axis between the CentileBrain-derived brain-PAD (A and C; -10 to +10 years) and ENIGMA-derived brain-PAD (B and D; - 20 to +20 years).

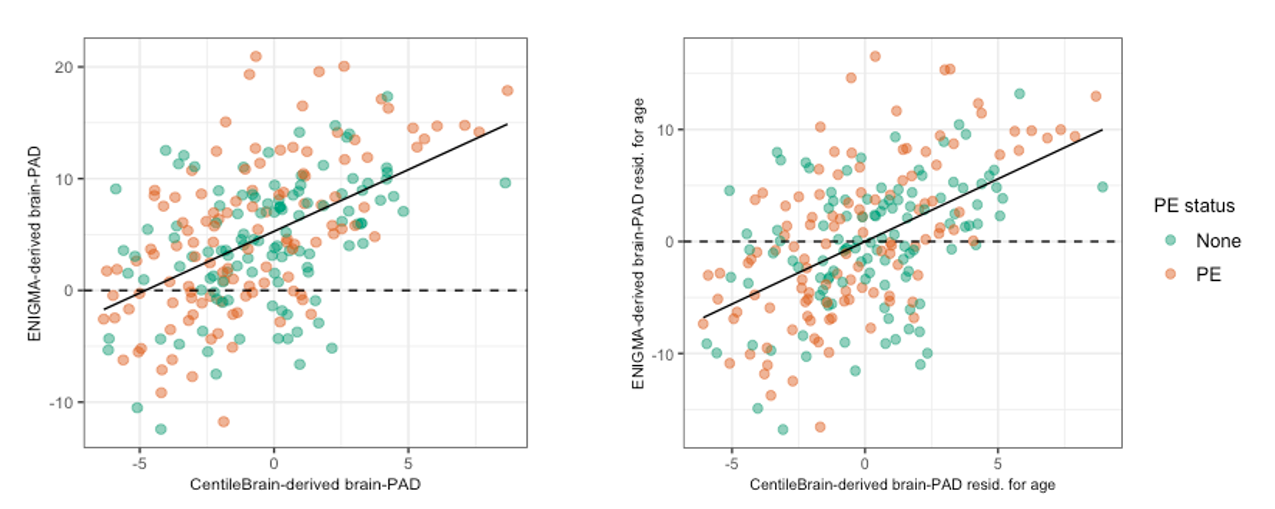

**Supplementary Figure S3. Pearson’s (r) and Spearman’s (rho) correlation between ENIGMA model-derived brain-PAD and CentileBrain model-derived brain-PAD in the current sample.** Left: A moderate correlation was observed between ENIGMA-derived brain-PAD and CentileBrain-derived brain-PAD (r = 0.52, p = 2 x 10^-16^; rho = 0.50). Right: The association was robust to age-related bias correction of the two brain-PAD measures (r = 0.52, p = 2 x 10^-16^; rho = 0.50), that is equivalent to a partial correlation adjusting for age.

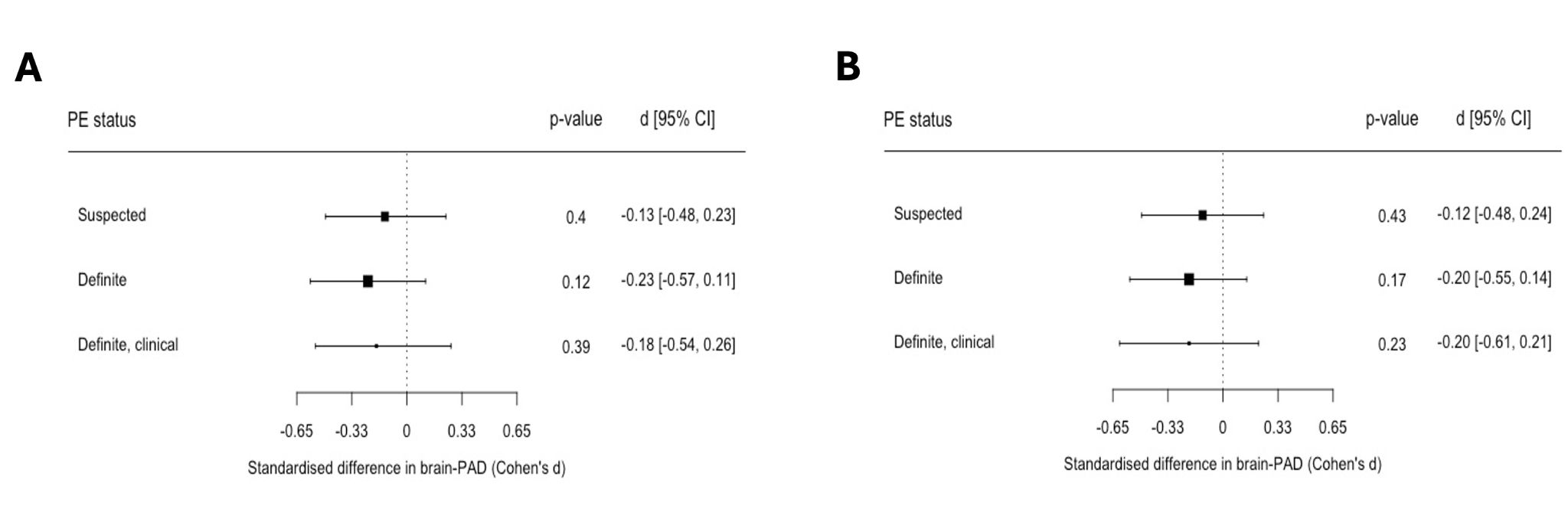

**Supplementary Figure S4. Difference in brain-PAD between suspected, definite, or definite, clinical PEs and those without PEs (reference).** (A) Forrest plot depicting the standardised difference (Cohen’s d and its 95% CI) in CentileBrain-derived brain-PAD between each PE sub-group and those without PEs. (B). Same as (A), but after outlier exclusion (n=2).

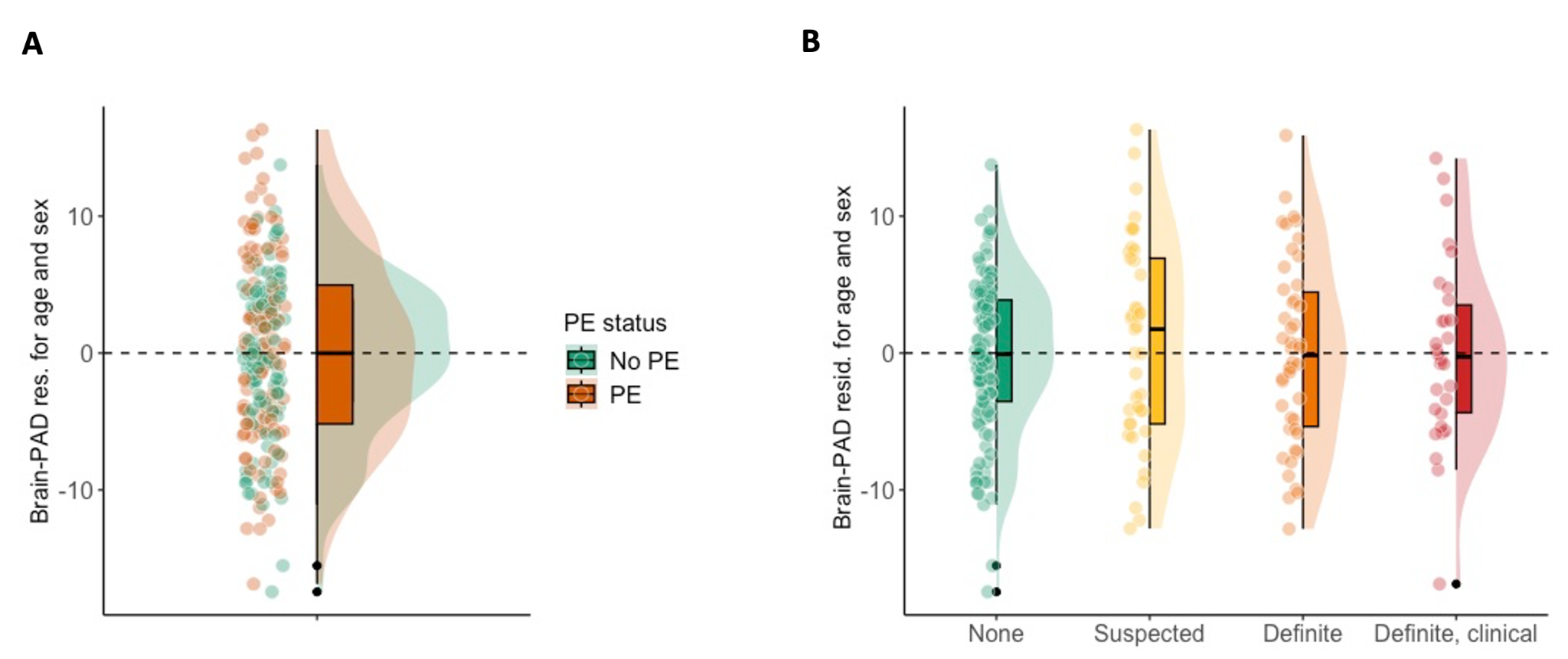

**Supplementary Figure S5.** Psychotic experiences and ENIGMA-derived brain-PAD. **(A)** brain-PAD estimates among participants without and with PE. **(B)** Brain-PAD estimates among those without PE (none) and each PE sub-category (suspected > definite > definite, clinical). Brain-PAD estimates are residualised for age and sex. Raincloud (half-density) plots depict group median, interquartile range, and potential outliers. Results of group comparison with ENIGMA-derived brain-PAD are summarised on Table S3 and Table S4.
